# The mental health impacts of child marriage in Ethiopia, India, Peru, and Vietnam: a longitudinal investigation

**DOI:** 10.64898/2026.08.03.26359570

**Authors:** Jasmine Lee, Thomas Steare, Jadhav Chakradhar, Revathi Ellanki, Jacques Wels, Praveetha Patalay

**Affiliations:** Centre for Longitudinal Studies, Social Research Institute, University College London, London, United Kingdom; Unit for Lifelong Health and Ageing, University College London, London, United Kingdom; Centre for Economic and Social Studies, Hyderabad, India; Health & Society Research Unit, Université libre de Bruxelles, Belgium

**Keywords:** psychological distress, wellbeing, gender equality, cohabitation, gender-based violence

## Abstract

**Background:** Child marriage/cohabitation before 18 years of age disproportionately affects girls worldwide, but its impacts on mental health are poorly understood. Existing research is mostly cross-sectional, hence unable to estimate causal impacts or longer-term outcomes of child marriage.

**Methods:** We analysed longitudinal cohort data across Ethiopia, India, Peru, and Vietnam from *Young Lives* to investigate the relationship between child marriage/cohabitation and life satisfaction and emotional symptoms. We used linear regression, difference-in-differences estimation (DiD), and propensity score matching (PSM) to examine short and longer-term impacts from age 19 to 29.

**Findings:** Across countries 5% (Peru) to 20% (India) of girls were child brides. Child marriage/cohabitation predicted lower life satisfaction – pooled estimates at age 19 (-0.25 [95%CI=0.40,-0.10]), across 19-29 (-0.22 [95%CI=-0.32,-0.13]), and heterogeneity estimates suggest consistently negative impacts across countries. However, child brides have lower life satisfaction at age 8 even before marriage occurs, likely due to pre-existing disadvantages such as poverty that increase child marriage likelihood. DiD and PSM analyses confirm that most of the differences are due to pre-existing vulnerability and there is a small additional impact of child marriage on worse life satisfaction at age 19 (pooled matched estimate -0.09 [95%CI=-0.19, 0.00]). No differences in emotional symptoms were observed, nor any differences by age of marriage within the child-married sample.

**Interpretation:** Child brides have lower life satisfaction from childhood through to early adulthood, which is mostly explained by prior disadvantages and risk factors, with a small additional impact of child marriage itself.

**Funding:** Wellcome Trust

## Introduction

Child marriage, defined as marriage or union under the age of 18 years, is considered a human rights violation and form of gender-based violence that disproportionately affects girls worldwide. Globally, the highest prevalence is in Sub-Saharan Africa (37%) and South Asia (30%). Although worldwide rates have declined over the last decade from 23% to 19% through policy and advocacy, it is estimated that full eradication of child marriage will take another 300 years at current pace.^1,2^ Girls from the poorest, rural households remain most at risk, especially in regions affected by the climate crisis, conflict, and rapid population growth.^3^

The impacts of child marriage on girls’ health, education, and economic outcomes are widely acknowledged. Two recent systematic reviews by Fan and Koski^4^ and Burgess^5^ focusing on the physical and mental health consequences found that depression, psychological distress, and suicidality were commonly linked to child marriage, driven by factors such as intimate partner violence (IPV), poverty, and childbirth challenges. However, all included studies with mental health outcomes were cross-sectional, where we cannot know whether child marriage preceded or caused poor mental health. This is relevant because girls at risk of child marriage are often already disadvantaged – from poorer, less educated, rural households, which are the same factors that also affect mental health.^6^ Only one recent study has examined the impacts of child marriage on life satisfaction with longitudinal data^6^ and found that child brides in India had lower life satisfaction even before marriage and up to age 22, with no negative effect of early marriage reported. In summary, existing evidence on the mental health impacts of child marriage is heterogeneous and limited,^4^ and the causal relationship between child marriage and mental health remains poorly understood. No longitudinal study to-date has investigated symptoms of mental illness, hence, we focused in this paper on both indicators of mental wellbeing (life satisfaction) and mental ill-health (emotional symptoms).

We investigated the mental health impacts of child marriage with cohort data from the *Young Lives* longitudinal study across Ethiopia, India, Peru, and Vietnam. The four countries represent different cultural, economic, and policy contexts across Sub-Saharan Africa, South Asia, Southeast Asia, and Latin America, with varying child marriage rates and contexts. India is home to one-third of the world’s child brides;^8^ there is substantial interstate variation in child marriage prevalence and *Young Lives* regions Andhra Pradesh and Telangana had slightly higher prevalence than the 2016 national average.^9^ In Ethiopia, four in ten young women were married or in union before 18, with rates significantly higher among rural and ethnic minority communities.^10^ In Peru, informal cohabitation (unión libre) is far more prevalent than formal marriage, often beginning due to pregnancy among girls from poorer families.^11^ In Vietnam, while national rates are lower, child marriage is concentrated among ethnic minority communities, with rates as high as 33% among H’mong people.^12^

We examined the impact of child marriage/cohabitation on mental health (life satisfaction and emotional symptoms) among girls married before 18 years using longitudinal regression with confounder adjustment, difference-in-differences estimation, as well as propensity score matching to account for pre-existing disadvantage and strengthen causal inference.

## Methods

### Participants

We used data from *Young Lives* (YL), a longitudinal cohort study which has followed more than 12,000 children from two birth cohorts (born in 1994/5, referred to as the 1994-cohort; or 2001/2, the 2001-cohort) over 25 years across Ethiopia, India (Andhra Pradesh and Telangana states), Peru, and Vietnam. The most recent round was in 2023 (except Vietnam, 2020-21). YL contains multi-dimensional data on development, education, and health across ages.^14^ It over-samples poorer families and findings should be interpreted accordingly.^13^ YL received ethics committee approval from relevant institutions, and participants provided informed consent for data collection.

Our analysis included girls only as child marriage is mainly experienced by girls, and few boys were married before 18 years (Ethiopia n=40, India n=6, Peru n=18, Vietnam n=45). All girls attending the first wave of data collection in 2002 were included with little attrition over time. 81% of original participants remained by 2023 across Ethiopia, India, and Peru, and 86% for Vietnam by 2021^14^; for more sample information and flowchart see Supplement S1.

### Measures

In the current study, in line with UNICEF’s definition, both marriage and cohabitation before age 18 constitute child marriage.^15^

The main exposure was being a child bride, which was derived from self-reported marital status (*“What is your current marital/cohabitation status?”*) and age of marriage (derived from date of marriage minus date of birth; also a secondary outcome), collected when the 1994-cohort was aged 19, 22 and 26, and when the 2001-cohort was aged 12, 15 and 19. Non-child-brides included girls who were not married or married after age 18. Variable details in Supplement S2.

Life satisfaction was measured with the self-reported Cantril ladder question,^16^ where participants rated their life on a nine-step ladder (1=worst possible life, 9=best possible life), collected at five timepoints for the 2001-cohort (ages 8, 12, 15, 19, 22) and six for the 1994-cohort (ages 12, 15, 19, 22, 26, 29), except in Vietnam (up to 19 and 26 respectively).

As a secondary outcome we included the self-reported Strengths and Difficulties Questionnaire emotional subscale (SDQ-E) (0-10), which measures internalising symptoms, and has been shown to be a reliable and valid measure of depression and anxiety (Supplement S3).^17^ This was only possible for the 1994-cohort at ages 15, 19, and 22 as SDQ-E was not included for the 2001-cohort.

We chose matching and confounding variables based on factors identified in previous research as household and individual characteristics related to child marriage.^18^ These included ethnicity/caste, religion, household characteristics at age 12 (urban/rural status, place in birth order, household size), socio-economic circumstances at age 12 (wealth index, housing quality index, consumer durables index, serious debt), education enrolment, involvement in paid work, aspirations for education, caregiver education, caregiver aspirations for the child’s education, and caregiver life satisfaction when child was aged 12. See Supplement S4 for measurement and inclusion justification.

### Statistical analysis

Three stages of analyses were conducted to answer different research questions and models were run for combined as well as separate cohorts (i.e. born 1994 and 2001) where possible. For each model there were three specifications: 1) adjusting for baseline (age 12) life satisfaction, 2) full covariate-adjusted models, and 3) propensity score matching (PSM) with full adjustment of covariates. All models accounted for clustering by sentinel site cluster. We applied PSM using 1:1 optimal matching in the *MatchIt* R package to minimise global distance and avoid sorting order bias,^19,20^ matching child brides and non-child brides (i.e., those never married or first married at aged 18 or older) with similar observed covariates to create a more comparable set of individuals. As a small number of individuals were married by age 12, meaning the exposure had already occurred when baseline life satisfaction was measured, we excluded these individuals in sensitivity analyses (post age-12 sensitivity check).

Models were run separately for each country first and where possible country estimates were pooled into a meta-analysis to generate an overall estimate and estimate heterogeneity (I^2^ statistic). This two-stage method allowed us to present country-specific as well as overall estimates, while examining whether effects are consistent across countries.

#### What is the impact of child marriage on mental health?

First, we conducted linear regression to predict differences in life satisfaction and emotional symptoms at age 19 between child brides and non-child-brides.

Second, to establish a more stringent causal inference for the effect of child marriage on life satisfaction changes between ages 12 and 19, we conducted a difference-in-differences (DiD) analysis. The model isolated the impact of child marriage on changes in wellbeing from ages 12 to 19 while controlling for pre-treatment differences and time-invariant unobserved confounders. Parallel trends assumption test (comparing life satisfaction trajectory prior to the baseline) was conducted in the 2001-cohort between ages 8 and 12 years (no life satisfaction measure available at age 8 in 1994-cohort). DiD was not possible with emotional symptoms as the earliest data was collected at age 15 when many girls had already married/cohabited.

#### What are the longer-term trajectories of mental health by child marriage status?

In order to determine whether child brides experience lower life satisfaction and more emotional symptoms across early adulthood (between ages 19 and 30), on average, compared to non-child brides, we used multi-level models to predict average outcomes across timepoints (ages 19, 22, 26, 29 for wellbeing; ages 19 and 22 for emotional symptoms). As data was collected only up to round 6 for Vietnam, models predicting average longer-term outcomes and trajectories were up to age 26, not age 29 in Vietnam.

As a further test of differences in life satisfaction trajectories in early adulthood (from age 19 to 30), we used multi-level models with an interaction term between child marriage status and age (19, 22, 26, 29) to examine whether the rate of change in life satisfaction differed by child marriage status. This analysis was conducted for the 1994-cohort only as the 2001-cohort was not old enough yet for longer-term trajectories.

#### Is there a difference in impact by age of marriage within child brides?

As secondary analyses, the same set of models were repeated with age of marriage as exposure, for the child bride subgroup only. This allowed us to examine whether the timing of marriage, not just the binary distinction of child marriage, impacted life satisfaction and emotional outcomes. DiD analysis compared life satisfaction between girls married by age 15 and those married between 16-17 years, allowing for a direct comparison of life satisfaction changes between ‘early- and later-treated’ groups. As a sensitivity check, similarly girls married by age 12 were excluded.

To address missing covariate data in the analytical sample (Supplement S5), multiple imputation with chained equations (*mice* package in R) was used. Imputations were performed separately for the 1994- and 2001-cohorts for each country with twenty imputations with a maximum of twenty iterations. Child marriage status and age of marriage were exposures and not imputed; covariates and outcomes were imputed where missing, with auxiliary variables included to improve imputations.

Statistical significance was set at p<0.05 for main effects, and due to greater power required for interaction models, p<0.10 was used as the threshold for interaction terms to reduce the risk of Type II errors.^21^ All variables included were measured with equivalent items across countries.

## Results

We analysed data from 5,711 girls: 1,440 from Ethiopia, 1,446 from India, 1,353 from Peru, and 1,472 from Vietnam.

Child marriage/cohabitation rates overall were highest in India (20.2%), followed by Ethiopia (9.7%), Vietnam (8.8%), and Peru (4.7%). Rates were higher in the 1994-cohort compared to the 2001-cohort across all countries. In Peru, cohabitation rather than formal marriage was the predominant union type. Age of marriage for child brides was lowest in Ethiopia, at 14.3 years (Supplement S6). Descriptive plots show that from age 8 to 29, child brides have lower life satisfaction overall in Ethiopia, India, and Vietnam, but no observed difference in emotional symptoms from age 15 to 22 (Figure 1).

**Figure 1.**
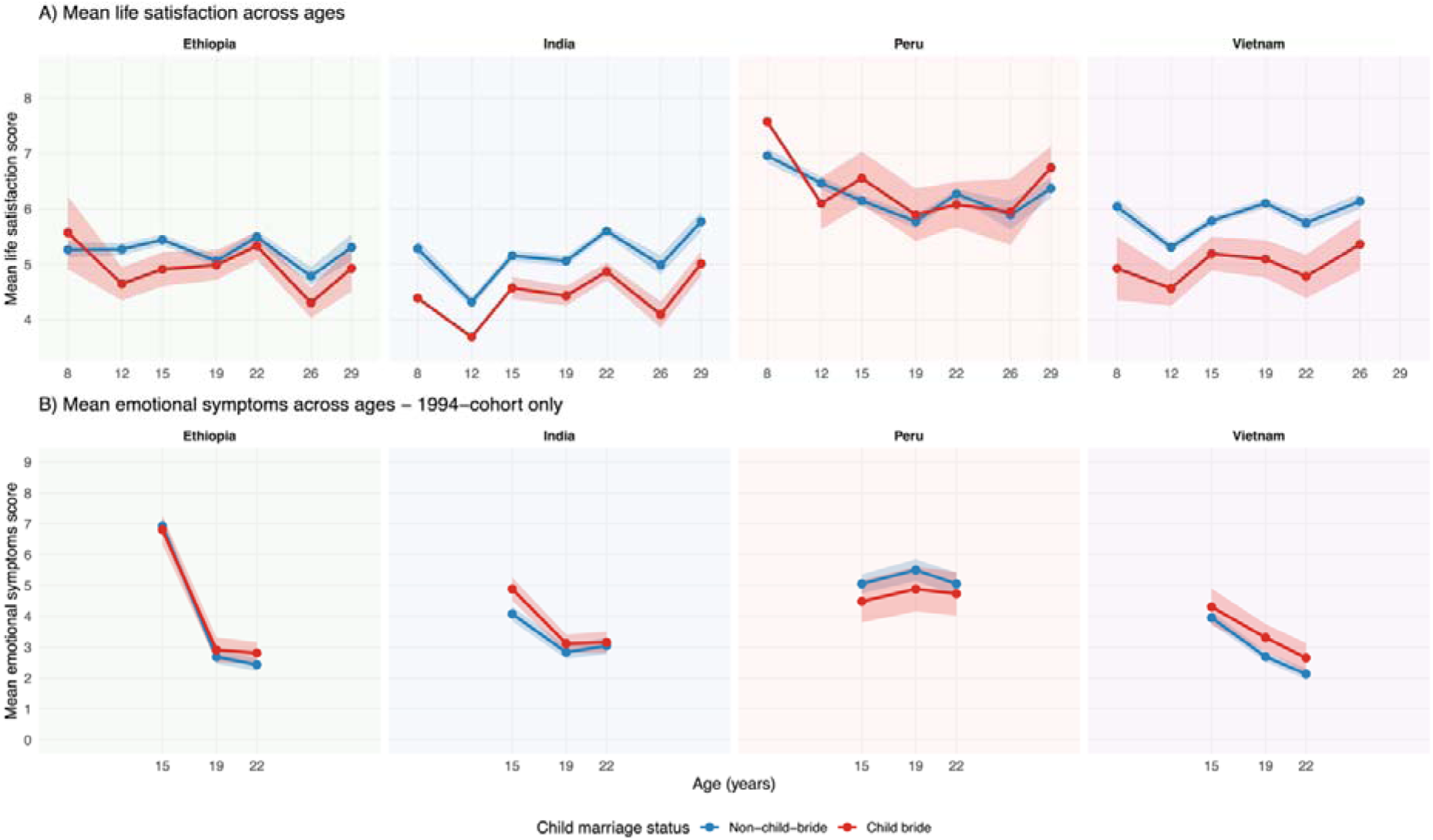
Trajectories of life satisfaction from 8 to 29 years (combined cohorts) and emotional symptoms from 15 to 22 years (1994-cohort only) by child marriage status across countries.

There was a higher proportion of child brides living in poorer families and rural areas, engaged in paid work by age 12, have lower aspirations for education, and caregivers with lower life satisfaction and no education, compared to non-child-brides. Table 1 presents demographic characteristics of the two groups across countries. For standardised mean differences and more detailed demographic information including ethnicity and religion, see Supplement S7.

**Table 1.**
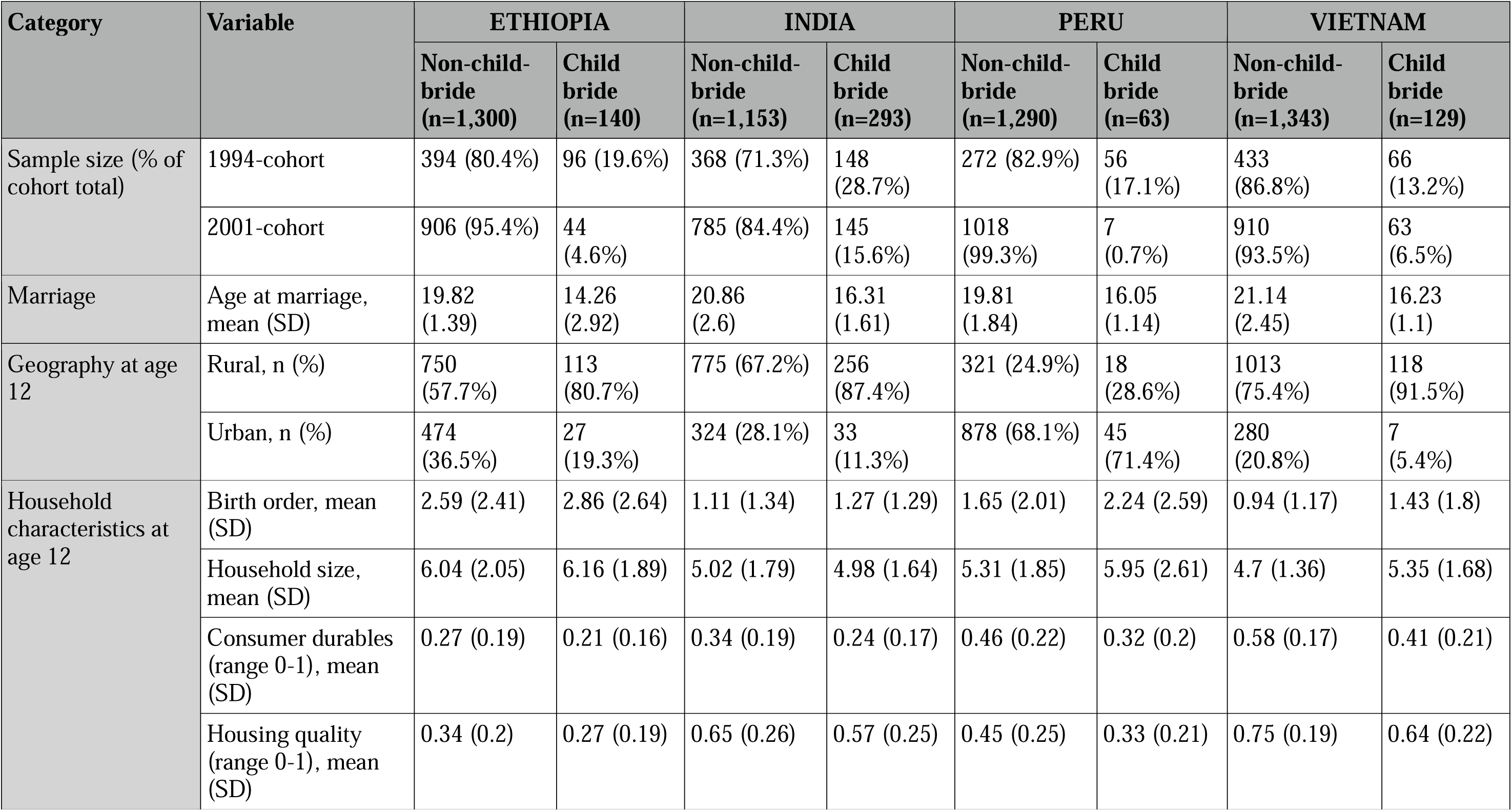
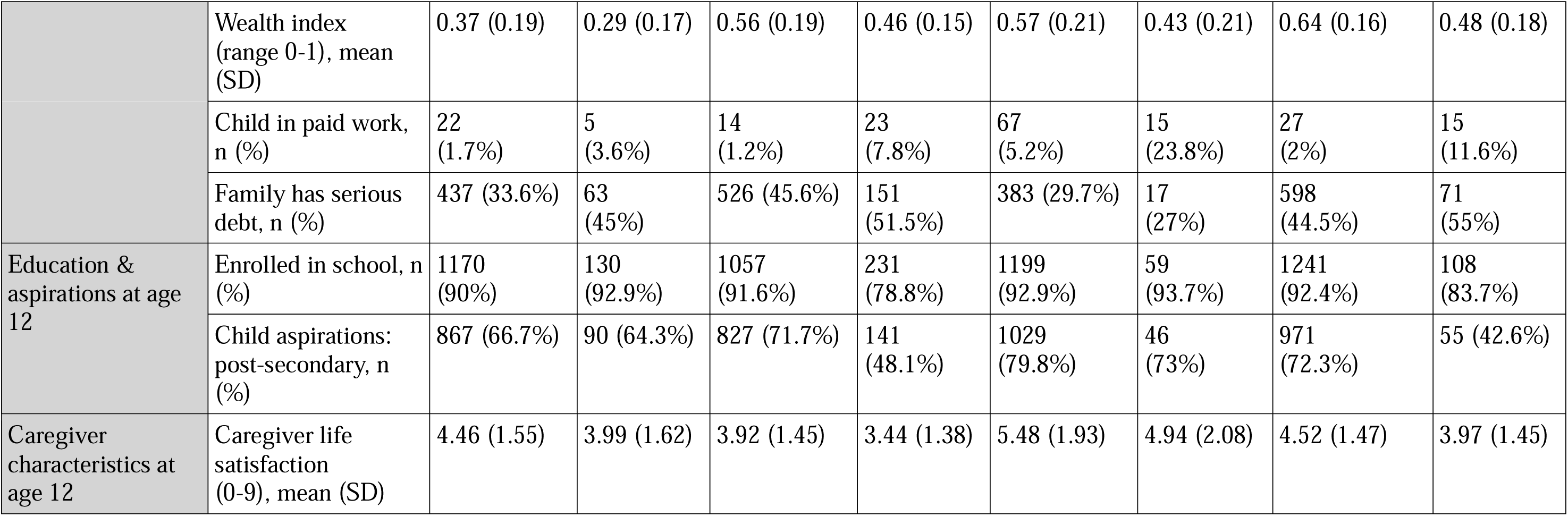
Demographic characteristics of child brides and non-child-brides.

### What is the impact of child marriage on mental health?

Overall, child brides had lower life satisfaction at age 19 (Figure 2) with a pooled estimate across countries of -0.25[CI -0.40, -0.10, I^2^=0.0]. Cohort-stratified analysis found a larger disadvantage in life satisfaction among the 1994-cohort child brides in India (-0.35[-0.69, - 0.01]) and Vietnam (-0.51[-0.96, -0.05]) than the 2001-cohorts (no observed difference), whereas in Ethiopia the disadvantage was clearer in the 2001-cohort (-0.73[-1.36, -0.10]) (Table 2).

**Figure 2.**
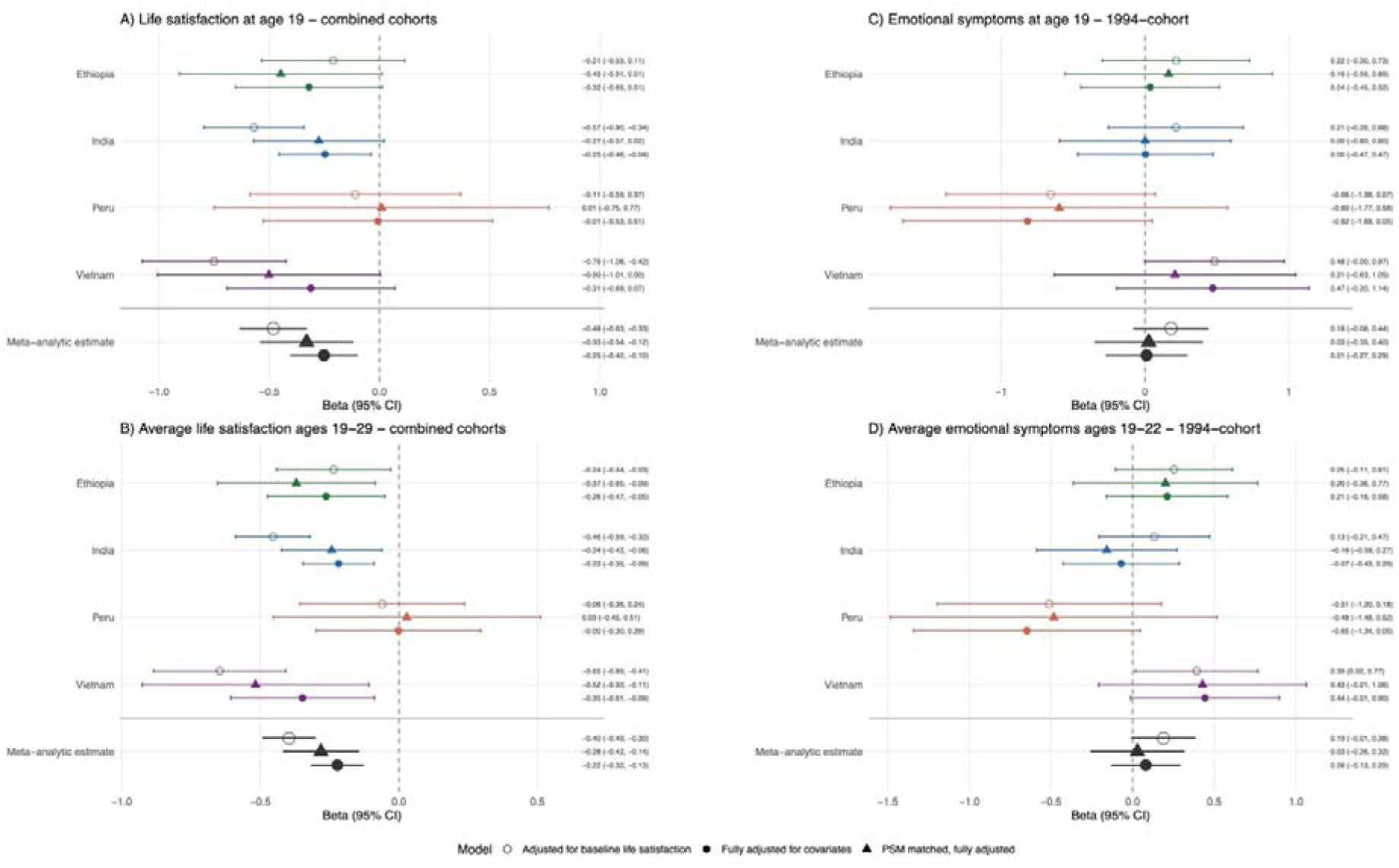
*Left:* Predicted life satisfaction at age 19 (A) and average life satisfaction across ages 19-29 (B) by child marriage status in combined cohorts across countries. *Right*: Predicted SDQ emotional symptoms at age 19 (C) and average emotional symptoms across ages 19-22 (D) by child marriage status in the 1994-cohort. Three levels of controls include adjustment for baseline life satisfaction, full adjustment of covariates, and propensity score matching (PSM) with full adjustment.

**Table 2.**
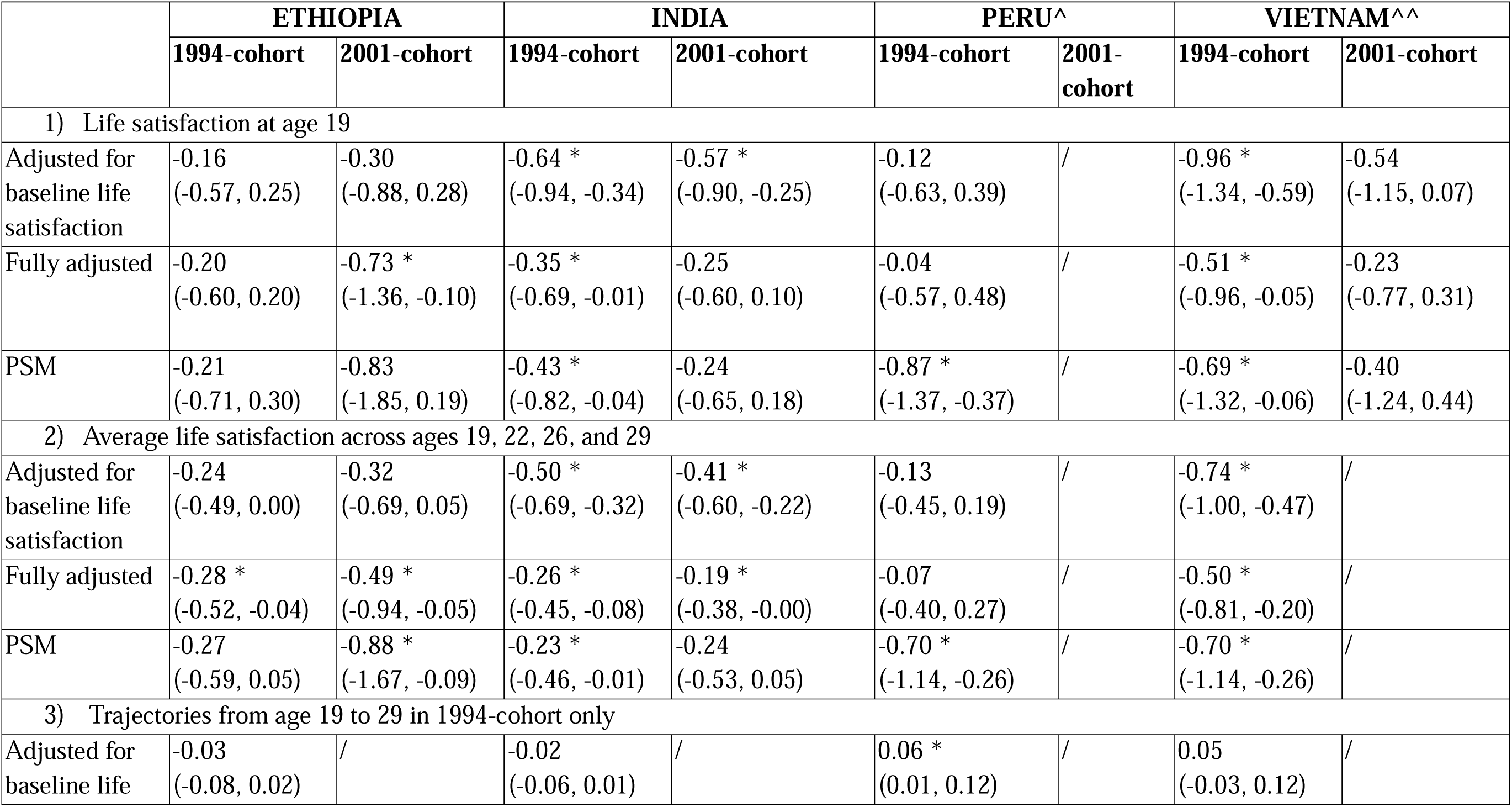
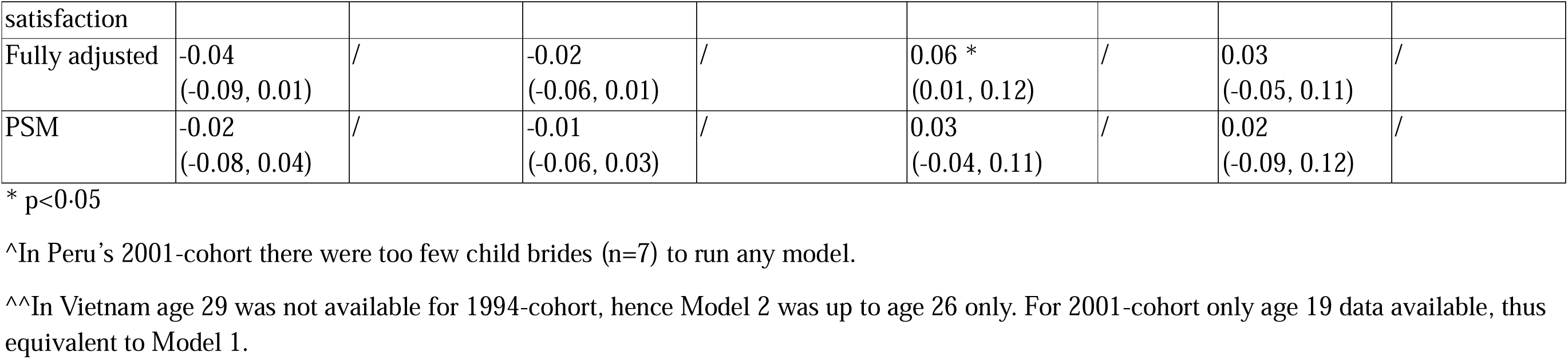
Summary of cohort-stratified models with child marriage status as exposure predicting child brides’: 1) life satisfaction at age 19; 2) Average life satisfaction across ages 19-29; 3) life satisfaction trajectories from ages 19-29. Non-child-brides were set as the reference level. Models either adjusted for baseline life satisfaction at age 12 only, fully adjusted for all covariates, or propensity score matched (PSM).

|  | ETHIOPIA |  | INDIA |  | PERU<sup>^</sup> |  | VIETNAM<sup>^^</sup> |  |
| --- | --- | --- | --- | --- | --- | --- | --- | --- |
|  | 1994-cohort | 2001-cohort | 1994-cohort | 2001-cohort | 1994-cohort | 2001-cohort | 1994-cohort | 2001-cohort |
| 1) Life satisfaction at age 19 |  |  |  |  |  |  |  |  |
| Adjusted for baseline life satisfaction | -0.16<br>(-0.57, 0.25) | -0.30<br>(-0.88, 0.28) | -0.64 *<br>(-0.94, -0.34) | -0.57 *<br>(-0.90, -0.25) | -0.12<br>(-0.63, 0.39) | / | -0.96 *<br>(-1.34, -0.59) | -0.54<br>(-1.15, 0.07) |
| Fully adjusted | -0.20<br>(-0.60, 0.20) | -0.73 *<br>(-1.36, -0.10) | -0.35 *<br>(-0.69, -0.01) | -0.25<br>(-0.60, 0.10) | -0.04<br>(-0.57, 0.48) | / | -0.51 *<br>(-0.96, -0.05) | -0.23<br>(-0.77, 0.31) |
| PSM | -0.21<br>(-0.71, 0.30) | -0.83<br>(-1.85, 0.19) | -0.43 *<br>(-0.82, -0.04) | -0.24<br>(-0.65, 0.18) | -0.87 *<br>(-1.37, -0.37) | / | -0.69 *<br>(-1.32, -0.06) | -0.40<br>(-1.24, 0.44) |
| 2) Average life satisfaction across ages 19, 22, 26, and 29 |  |  |  |  |  |  |  |  |
| Adjusted for baseline life satisfaction | -0.24<br>(-0.49, 0.00) | -0.32<br>(-0.69, 0.05) | -0.50 *<br>(-0.69, -0.32) | -0.41 *<br>(-0.60, -0.22) | -0.13<br>(-0.45, 0.19) | / | -0.74 *<br>(-1.00, -0.47) | / |
| Fully adjusted | -0.28 *<br>(-0.52, -0.04) | -0.49 *<br>(-0.94, -0.05) | -0.26 *<br>(-0.45, -0.08) | -0.19 *<br>(-0.38, -0.00) | -0.07<br>(-0.40, 0.27) | / | -0.50 *<br>(-0.81, -0.20) | / |
| PSM | -0.27<br>(-0.59, 0.05) | -0.88 *<br>(-1.67, -0.09) | -0.23 *<br>(-0.46, -0.01) | -0.24<br>(-0.53, 0.05) | -0.70 *<br>(-1.14, -0.26) | / | -0.70 *<br>(-1.14, -0.26) | / |
| 3) Trajectories from age 19 to 29 in 1994-cohort only |  |  |  |  |  |  |  |  |
| Adjusted for baseline life | -0.03<br>(-0.08, 0.02) | / | -0.02<br>(-0.06, 0.01) | / | 0.06 *<br>(0.01, 0.12) | / | 0.05<br>(-0.03, 0.12) | / |
| satisfaction |  |  |  |  |  |  |  |  |
| Fully adjusted | -0.04<br>(-0.09, 0.01) | / | -0.02<br>(-0.06, 0.01) | / | 0.06 *<br>(0.01, 0.12) | / | 0.03<br>(-0.05, 0.11) | / |
| PSM | -0.02<br>(-0.08, 0.04) | / | -0.01<br>(-0.06, 0.03) | / | 0.03<br>(-0.04, 0.11) | / | 0.02<br>(-0.09, 0.12) | / |

\* $p < 0.05$
^In Peru's 2001-cohort there were too few child brides ( $n=7$ ) to run any model.
^^In Vietnam age 29 was not available for 1994-cohort, hence Model 2 was up to age 26 only. For 2001-cohort only age 19 data available, thus equivalent to Model 1.

As a sensitivity check, we excluded girls married/cohabited by age 12 (post age-12 sensitivity check, excluding Ethiopia n=26, India n=13, Peru n=1, Vietnam n=1). Results were consistent, with more negative effects in Ethiopia (-0.40[-0.75, -0.05]) (Supplement S8). In the PSM samples, child brides did not have lower life satisfaction in any country at age 19 (Figure 2; matched sample sizes in Supplement S9; Peru 2001-cohort not matched as n=7).

Difference-in-differences (DiD) estimation indicated no overall affect in the adjusted regression, but there was evidence of a pooled effect in the PSM samples (-0.09[-0.19, 0.00] with I^2^=1.7%) (Figure 3; Supplement S10), suggesting a small negative causal effect of child marriage on life satisfaction beyond pre-existing disadvantages across countries, with little cross-country heterogeneity. Post age-12 sensitivity checks yielded similar results (Supplement S8).

**Figure 3.**
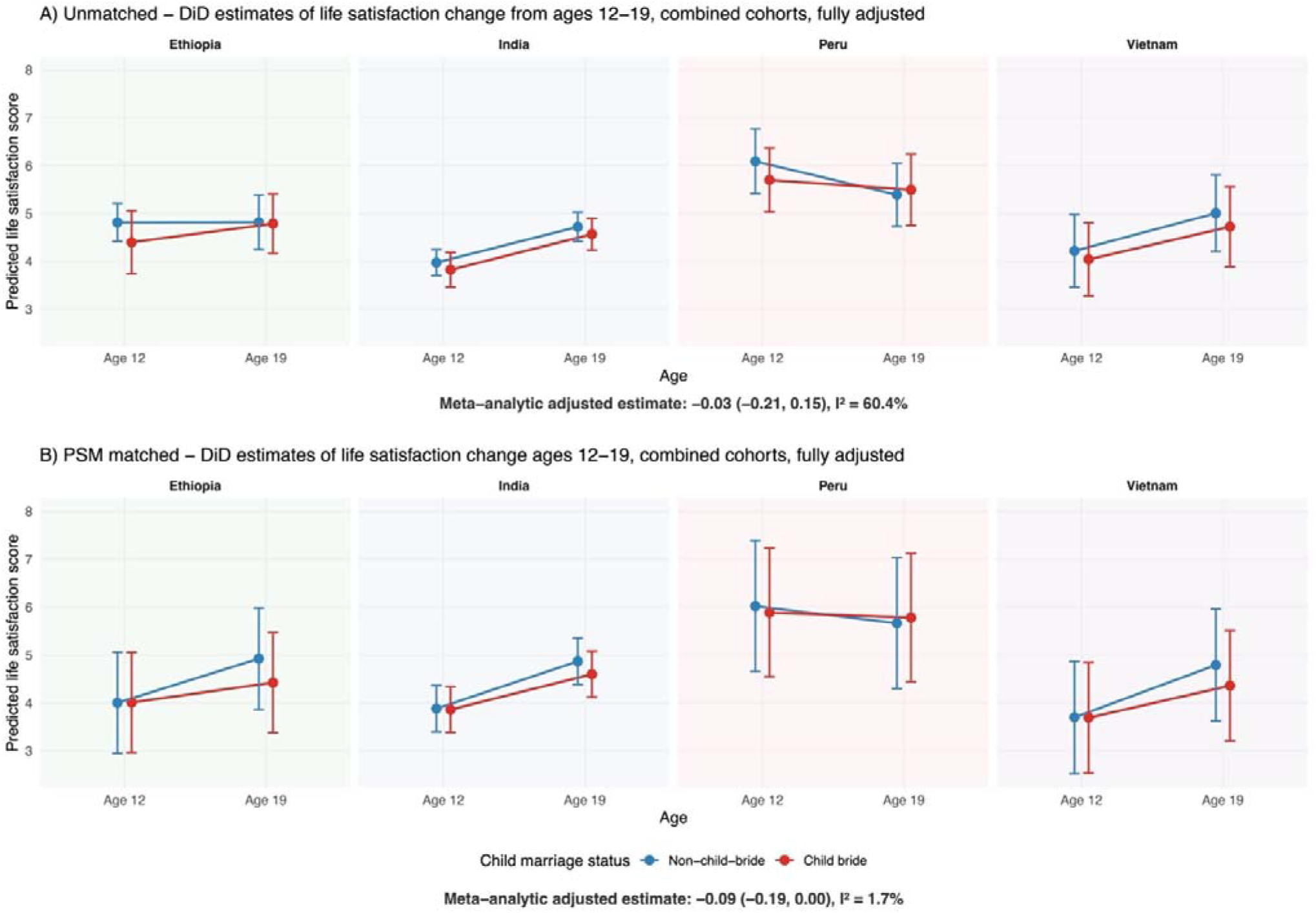
Difference-in-differences (DiD) estimated marginal means of life satisfaction from age 12 to 19 in covariate-adjusted unmatched (A) and propensity score matched (PSM) (B) combined cohorts across countries.

Child brides did not appear to have greater emotional symptoms in any country after covariate adjustment or matching (Figure 2).

### What are the longer-term trajectories of mental health by child marriage status?

Lower life satisfaction was reported on average among child brides across ages 19, 22, 26 and 29; the covariate-adjusted pooled estimate was -0.22[-0.32, -0.13], I^2^=5% and the matched pooled estimate was -0.28[-0.42, -0.14], I^2^=12.0% (Figure 2). Coefficients were higher in Vietnam, Ethiopia, and India compared to Peru, in both overall and cohort-stratified models (Table 2), however, low heterogeneity in the pooled estimate indicates this might not reflect systematic country differences.

Analyses of life satisfaction trajectories from age 19 to 29 in the 1994-cohort found no trajectory differences between child brides and non-child-brides in Ethiopia, India, nor Vietnam (Table 2; Supplement S11). In Peru, however, the life satisfaction of child brides improved 0.06[0.01, 0.12] points per year more than non-child-brides, with similar results in post age-12 sensitivity checks (Supplement S8). However, in matched analyses, this was not replicated and no trajectory differences were observed (Table 2).

Analysis of average emotional symptoms (SDQ-E) across ages 19 and 22 years in the 1994-cohort indicated no differences in both unmatched and matched samples (Figure 2).

### Is there a difference in impact by age of marriage within child brides?

The sample was largely reduced as analyses were restricted to only child brides who reported age of marriage/cohabitation (Ethiopia n=99, India n=293, Peru n=63, Vietnam n=115), thus underpowered to detect small effects. We found few differences in outcomes between those married earlier and later across models, outcomes, and specifications (Supplement S12).

## Discussion

Among the *Young Lives* longitudinal cohorts of girls across Ethiopia, India (Andhra Pradesh and Telangana), Peru, and Vietnam, rates of child marriage/cohabitation were highest in India (20%), and across countries lower amongst those born in 2001 than 1994, potentially reflecting continuing efforts to end child marriage globally.^22^ Our findings highlighted that life satisfaction gaps in childhood are present years before child marriage occurs, highlighting prior disadvantage and vulnerability for this group. Across all countries the child married/cohabited group was more likely to be poorer, living rurally, and have lower aspirations for education. Causally identified models suggested that there was a small additional causal impact of child marriage on lower life satisfaction at age 19.

We included two outcomes in this study, life satisfaction and emotional symptoms; the latter only available for a narrower age range and the 1994-cohort. Where comparable, we found different patterns of results for each outcome. Child brides experienced lower life satisfaction from age 8 through to age 29 compared to non-child-brides – suggesting they were already more vulnerable even before marriage/cohabitation, likely due to sociodemographic circumstances that increased both the risk of child marriage, such as poverty, and the risk of worse wellbeing/mental health. This was further confirmed by matched analysis where comparing similar child brides and non-child brides showed no differences in life satisfaction trajectories over time. In contrast, no differences in emotional symptoms were observed across the two groups in any country, which was confirmed by formal analysis. This is in contrast to cross-sectional studies that have reported gaps in distress between child brides and non-child brides in Ethiopia.^23^ This may reflect the more global nature of the life satisfaction measure which captures an overall evaluation of all aspects of life, rather than a narrow set of depression and anxiety symptoms captured by the emotional symptoms measure. Differences in more severe mental health outcomes such as suicidal thoughts and attempts have been reported in Ethiopia,^24^ however, research remains limited in this area, with no other cross-sectional or longitudinal studies for severe outcomes identified across these countries.^5^

Our analyses found that child brides have lower life satisfaction in late adolescence and through to early adulthood in Ethiopia, India, and Vietnam. This effect was not identified in Peru, likely due to small sample sizes as only 63 girls were married/cohabited before 18. It may also reflect the predominant type of union, as cohabitation is far more common than formal marriage in Peru and may relate differently to wellbeing. For example, cohabitation may be seen by girls as a chance to improve their own well-being, despite reduced educational opportunities and increased risk of intimate partner violence.^11^ Differences in type of union may be a source of cross-country heterogeneity, however, the low statistical heterogeneity of estimates between countries suggests that any cross-country variation in observed estimates is by chance, and that the association of child marriage with later life satisfaction is similarly negative across these four countries.

More causally informative analyses trying to isolate the impacts of child marriage using difference-in-differences and propensity score matching found small impacts on wellbeing. This extends previous research from Kanji et al.^7^ who used only the 1994-cohort of girls in India (n=486, including 139 child brides) and reported no causal effect of child marriage on wellbeing (0.32[-0.17, 0.81]) in the matched sample – which includes our pooled estimate of -0.09. Our larger pooled matched sample (n=1,236 across four countries, including 618 child brides) yields greater precision, allowing us to detect a small negative effect that a smaller sample may not. Nevertheless, it is worth noting that the magnitude of this effect is small (0.1 point lower on a 1-9 scale of life satisfaction).

We found no evidence within child brides that marriage/cohabitation at an earlier age was associated with worse mental health, though this may be attributed to analyses being restricted within the child marriage group and hence with limited power to detect small effects. Any differences by age of child marriage may also be difficult to disentangle from pre-existing disadvantages that previous research has found to predispose girls to be married earlier in these contexts – such as poverty and lower levels of education,^6^ which are adjusted for in our analyses.

While the overall effects of child marriage/cohabitation were negative across countries, girls from different backgrounds and communities may have substantially different experiences of child marriage. Potential mechanisms include early pregnancy and childbirth complications, physical and sexual violence,^25^ reduced educational and employment prospects,^26^ and restricted agency and control over resources.^27^ Some qualitative research also suggests reasons for improvements in wellbeing linked to economic benefits, such as financial stability for girls facing extreme poverty.^28^ Child marriage customs vary considerably across ethnic groups, communities, and regions even within the same country.^28^ Our paper focuses on understanding population-level average impacts, but we recognise that these average estimates will mask heterogeneity in experience and outcomes within these populations. Future research is also needed to understand how consequences of child marriage, such as early pregnancy and school dropout, mediate its impact on mental health.

To our knowledge, this is the first cross-national longitudinal study examining the short- and longer-term impacts of child marriage/cohabitation on mental health. Analyses of longitudinal data across four countries enabled comparison across diverse contexts and testing of generalisability. Strong prior information on sociodemographic characteristics (such as rural residence and education enrolment) as well as psychological characteristics (such as child’s aspirations and parent’s aspirations for child’s education) allowed for comprehensive adjustment in our analytical models to isolate the effect of child marriage/cohabitation. For difference-in-differences models, we could not run the parallel trends assumption test for the 1994-cohort as no data was collected before age 12.

## Conclusions

Overall, our findings highlight that the relationship between child marriage and mental health is complex. Contrary to evidence from cross-sectional associations reported in past studies,^5^ our findings suggest child marriage/cohabitation is likely a downstream consequence and extension of early childhood disadvantages that contribute to poor mental health, rather than a primary driver in itself.

To help improve population mental health, interventions to reduce rates of child marriage should focus on the structural circumstances leading to both poor mental health and early marriage/cohabitation^5^, such as cash or in-kind transfers to support girls’ school attendance, particularly in ultra-poor households – which evaluations show to be the most successful interventions in delaying marriage,^2^ rather than relying only on policy means that do not address underlying causes. Beyond financial transfers, there is evidence that community-based programmes targeting social drivers, gender rights, and livelihoods can reduce rates of child marriage,^29^ and the mental health impacts of such programmes should also be considered. Importantly, improving the implementation of existing programmes may matter as much as introducing new ones: in India, for example, several initiatives aim to keep girls in school and delay marriage, and relevant laws on child marriage and dowry exist, but their success often depends on effective implementation.^30^

Alongside tackling the embedded social, cultural, and economic causes of child marriage to improve outcomes for girls, it is important to recognise that child marriage/cohabitation is still widely occurring, and our findings highlight the worse long-term life satisfaction for this group, for whom accessible and acceptable forms of support should be available. Preventing child marriage should not be an isolated effort, but part of a broader agenda that invests in girls’ education, poverty reduction, and mental health services.

## Data Availability

The data used in this study are available through the UK Data Service.

## Acknowledgements

We would like to thank the Young Lives families and research teams for the use of these data, and especially the staff and families we visited in Hyderabad, India, for their generous hospitality.

JL is funded by a University College London-Wellcome Trust doctoral training fellowship in mental health science (218497/Z/19/Z). The analysis and interpretation of these data has been done solely by the authors, and the findings or views reported in this paper should not be attributed to any of the funders or data providers.

## Author contributions

Conceptualisation: PP, RE, JC, JL; Methodology: PP, JL, RE, JC, TS; Analysis: JL; Code verification: TS, JW; Writing- drafting: JL, PP; Writing- revision and editing: All authors

## Data sharing

The data used in this study are available through the UK Data Service.

## Abbreviations

CB: Child brides
CI: Confidence interval
DiD: Difference-in-differences
IPV: Intimate partner violence
NA: Not applicable
NCB: Non-child-brides
PSM: Propensity score matching
SDQ-E: Strengths and Difficulties Questionnaire emotional subscale
YL: Young Lives

## Notes

### Competing Interest Statement

The authors have declared no competing interest.

## Bibliography

1 UNICEF. Is an end to child marriage within reach? Latest trends and future prospects. 2023 update. UNICEF DATA. 2023. https://data.unicef.org/resources/is-an-end-to-child-marriage-within-reach/ (accessed Jan 22, 2025).

2 Malhotra A, Elnakib S. 20 years of the evidence base on what works to prevent child marriage: a systematic review. J Adolesc Health 2021; 68: 847–62.

3 CRANK. Evidence review: Child marriage interventions and research from 2020 to 2022. Child Marriage Research to Action Network (the CRANK), 2023 https://www.girlsnotbrides.org/documents/1904/CRANK_Evidence_review_Child_marriage_interventionshttps://www.girlsnotbrides.org/en/documents/1904/CRANK_Evidence_review_Child_marriage_interventions__research_2020-22.pdf (accessed Jan 22, 2025).

4 Fan S, Koski A. The health consequences of child marriage: a systematic review of the evidence. BMC Public Health 2022; 22: 309.

5 Burgess RA, Jeffery M, Odero SA, Rose-Clarke K, Devakumar D. Overlooked and unaddressed: a narrative review of mental health consequences of child marriages. PLOS Glob Public Health 2022; 2: e0000131.

6 Briones K, Porter C. How does teenage marriage and motherhood affect the lives of young women in Ethiopia, India, Peru and Vietnam? Oxford: Young Lives, 2019.

7 Kanji S, Carmichael F, Darko C, Egyei R, Vasilakos N. The impact of early marriage on the life satisfaction, education and subjective health of young women in India: a longitudinal analysis. J Dev Stud 2024; 60: 705– 23.

8 UNICEF. Ending child marriage: a profile of progress in India. UNICEF DATA. 2023. https://data.unicef.org/resources/ending-child-marriage-a-profile-of-progress-in-india-2023/ (accessed Jan 22, 2025).

9 Gausman J, Kim R, Kumar A, Ravi S, Subramanian SV. Prevalence of girl and boy child marriage across states and Union Territories in India, 1993–2021: a repeated cross-sectional study. Lancet Glob Health 2024; 12: e271–81.

10 UNICEF Ethiopia. Understanding the situation of children. https://www.unicef.org/ethiopia/understanding-situation-children (accessed May 20, 2026).

11 Rojas V, Bravo F. Experiences of cohabitation, marriage and parenting in Peruvian adolescents and youth. Oxford: Young Lives, 2020.

12 UNFPA Vietnam. Ending child marriage: towards a world where girls are free to dream. https://vietnam.unfpa.org/en/news/ending-child-marriage-towards-world-where-girls-are-free-dream (accessed May 20, 2026).

13 Young Lives. Sampling and attrition. https://www.younglives.org.uk/sampling-and-attrition (accessed July 27, 2026).

14 Favara M, De Los Ángeles Molina M, Sánchez A, et al. Cohort profile update: the Young Lives study. Int J Epidemiol 2026; 55: dyag087.

15 UNICEF. Child marriage. UNICEF DATA. 2023. https://data.unicef.org/topic/child-protection/child-marriage/ (accessed Jan 22, 2025).

16 Levin KA, Currie C. Reliability and validity of an adapted version of the Cantril Ladder for use with adolescent samples. Soc Indic Res 2014; 119: 1047–63.

17 Armitage JM, Tseliou F, Riglin L, et al. Validation of the Strengths and Difficulties Questionnaire (SDQ) emotional subscale in assessing depression and anxiety across development. PLoS One 2023; 18: e0288882.

18 Singh R, Vennam U. Factors shaping trajectories to child and early marriage: evidence from Young Lives in India. 2016 https://www.younglives.org.uk/sites/default/files/migrated/YL-WP149-Trajectories%20to%20early%20Marriage.pdf (accessed Feb 17, 2025).

19 Ho D, Imai K, King G, Stuart EA. MatchIt: nonparametric preprocessing for parametric causal inference. J Stat Softw 2011; 42: 1–28.

20 Gu XS, Rosenbaum PR. Comparison of multivariate matching methods: structures, distances, and algorithms. J Comput Graph Stat 1993; 2: 405–20.

21 Thiese MS, Ronna B, Ott U. P value interpretations and considerations. J Thorac Dis 2016; 8: E928–31.

22 UNICEF. 25 million child marriages prevented in last decade due to accelerated progress, according to new UNICEF estimates. 2018. https://www.unicef.org/turkiye/en/press-releases/25-million-child-marriages-prevented-last-decade-due-accelerated-progress-according (accessed June 18, 2026).

23 John NA, Edmeades J, Murithi L. Child marriage and psychological well-being in Niger and Ethiopia. BMC Public Health 2019; 19: 1029.

24 Gage AJ. Association of child marriage with suicidal thoughts and attempts among adolescent girls in Ethiopia. J Adolesc Health 2013; 52: 654–6.

25 Irani M, Latifnejad Roudsari R. Reproductive and sexual health consequences of child marriage: a review of literature. J Midwifery Reprod Health 2019; 7: 1491–7.

26 Wodon Q, Savadogo A, Yedan A, et al. Economic impacts of child marriage: global synthesis report. Washington, DC: World Bank, 2017.

27 Tauseef S, Sufian FD. The causal effect of early marriage on women’s bargaining power: evidence from Bangladesh. World Bank Econ Rev 2024; 38: 598–624.

28 Ahonsi B, Fuseini K, Nai D, et al. Child marriage in Ghana: evidence from a multi-method study. BMC Women’s Health 2019; 19: 126.

29 Amin S, Saha JS, Ahmed JA. Skills-building programs to reduce child marriage in Bangladesh: a randomized controlled trial. J Adolesc Health 2018; 63: 293–300.

30 Jejeebhoy SJ. Ending child marriage in India: drivers and strategies. UNICEF, 2019 https://www.unicef.org/india/media/2556/file/Drivers-strategies-for-ending-child-marriage.pdf (accessed July 28, 2026).

