## Supplement for "The mental health impacts of child marriage in Ethiopia, India, Peru, and Vietnam: a longitudinal investigation"

### S1. Flowchart and cohort information

A) Information on Young Lives cohorts:

*Young Lives (YL)* comprises of two cohorts, the cohort born in 2001/2 (known as 2001-cohort in this paper, or Younger Cohort in YL documentation) and 1994/5 (1994-cohort in this paper, or Older Cohort).

Quantitative data has been collected over seven rounds between 2002 and 2023.

|  | | Round 1 (2002) | Round 2 (2006) | Round 3 (2009) | Round 4 (2013) | Round 5 (2016) | Round 6 (Phone Survey)  (2020-2021) | Round 7  (2023) |
| --- | --- | --- | --- | --- | --- | --- | --- | --- |
| 2001-cohort | Age | 1 | 5 | 8 | 12 | 15 | 19-20 | 22 |
|  | Rates | 100% | 96.7% | 95.8% | 93.2% | 93.2% | 87.7% | 83.5% |
| 1994-cohort | Age | 8 | 12 | 15 | 19 | 22 | 26-27 | 29 |
|  | Rates | 100% | 98.3% | 96.9% | 90.9% | 87.4% | 82.5% | 75.7% |

The Round 7 retention rate is calculated over three countries, Ethiopia, India, and Peru, as data collection was not conducted in Vietnam due to changes in international data-transfer laws. Attrition was lowest in India and highest in Ethiopia, likely due to the ongoing armed conflict in Tigray and Amhara. The study did not follow participants who migrated to other countries.

This information is based on the most recent cohort profile from the Young Lives team.^1^

B) Sample flowchart of main analyses

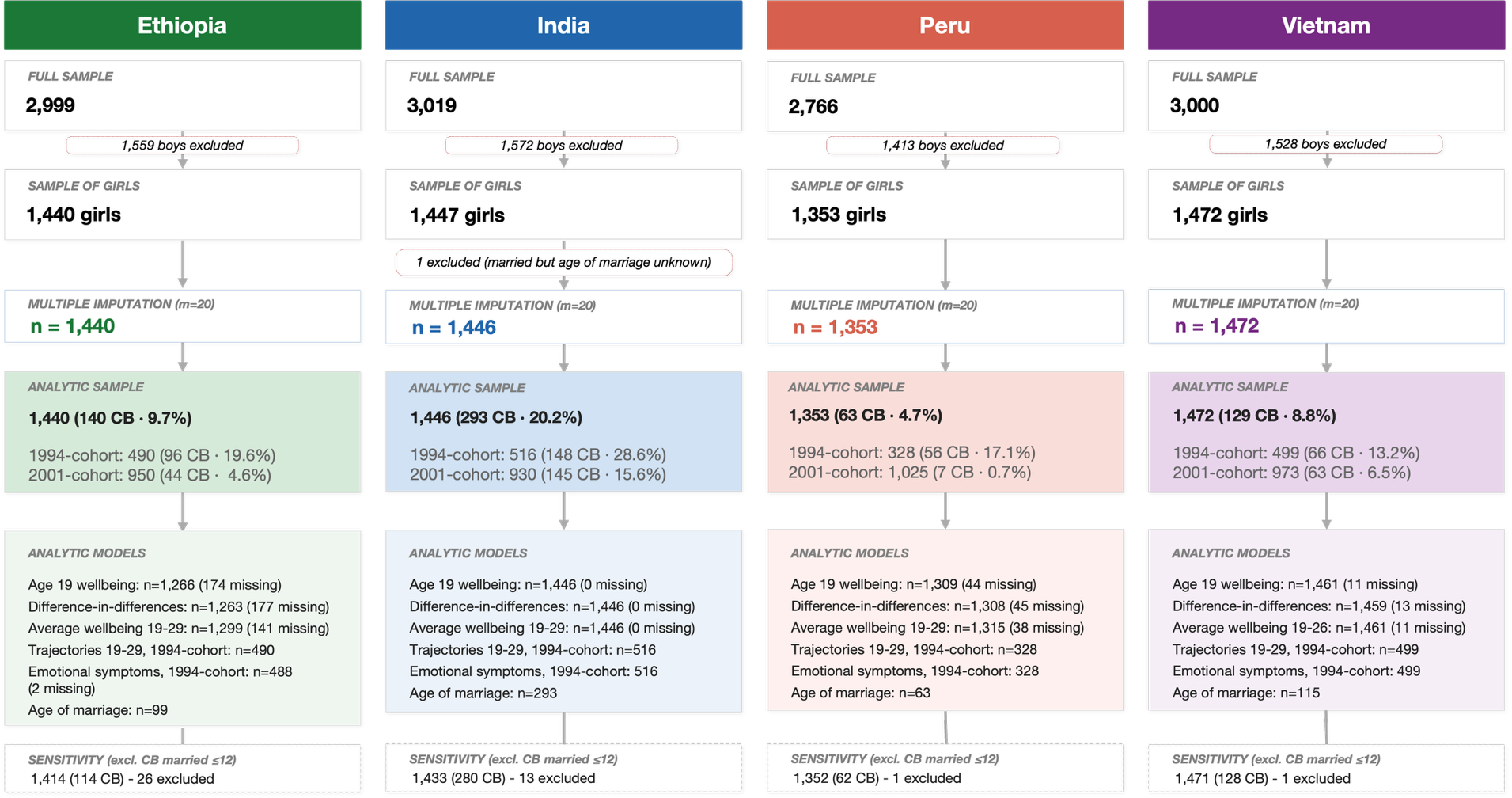

NCB: Non-child-bride; CB: Child bride

### S2. Constructing child marriage/cohabitation variables

Item for child marriage/cohabitation: “What is your current marital/cohabitation status?”

Data was collected in Rounds 4, 5, and 6 – the 1994-cohort was aged 19, 22, and 26; and 2001-cohort was aged 12, 15, and 19 respectively.

Response options for the item included:

- Single (never married)
- Married (to different person/ newly married since previous round)
- Cohabitant (with different person/ newly cohabiting since previous round)
- Widow(er)
- Divorced
- Separated
- Married to same spouse as in previous round
- Married since previous round to partner from previous round
- Cohabitant with the same partner as previous round.

Everyone who responded ‘Single’ was categorised as ‘never married/cohabited’, whereas all other responses were grouped as ‘ever married/cohabited’. Based on age of marriage/cohabitation (derived from date of marriage minus date of birth), those ‘ever married/cohabited’ were then categorised as a ‘child bride’ if marriage/cohabitation age was below 18.

As age of marriage had a lot of missing data, we also categorised individuals as child brides if they responded yes to any of the categories other than ‘Single’ when being interviewed in a round where they were under 18 years (i.e. 2001-cohort at age 12 or 15).

One individual in India from the 1994-cohort was married but missing age of marriage, thus unable to determine whether they were married before or after 18 and were excluded from the sample.

If there was discrepancy across rounds, as long as an individual had responded married/cohabited categories in one round they would be counted as child brides. The lowest age of marriage/cohabitation would be used if the ages recorded were different across rounds.

### S3. Strengths and Difficulties Questionnaire (SDQ) variables

We used the SDQ emotional subscale, which included the following 5 items:

1. ‘You worry a lot’
2. ‘You get a lot of headaches, stomach aches or sickness.’
3. ‘You are often unhappy, downhearted or tearful.’
4. ‘You are nervous in new situations’
5. ‘You have many fears, you are easily scared.’

The Young Lives scale was ‘Certainly true for you’, ‘A little true for you’, ‘Not true for you’ in Peru, which we reverse-coded to match on to conventional SDQ scoring of ‘Not true’ (0), ‘Somewhat true’ (1), and ‘Certainly true’ (2). In Ethiopia, India, and Vietnam the questions had 4 items – ‘Strongly disagree’, ‘Disagree’, ‘Agree’, ‘Strongly agree’, hence we collapsed ‘Strongly disagree’ and ‘Disagree’ into ‘Not true’, and renamed ‘Agree’ as ‘Somewhat true’, and ‘Strongly agree’ as ‘Certainly true’ to match SDQ scoring.

The range of score for each item is 0-2, and summing the five items produces an overall emotional score ranging 0-10.

### S4. Confounder list and justification

| **Confounder** | **Details** | **Age collected** |
| --- | --- | --- |
| Ethnicity/ caste | Child’s ethnic group or caste (India only) | All ages |
| Religion | Child’s religion | All ages |
| Place in birth order | The order the child is born into the family | Age 5 for 2001-cohort, age 8 for 1994-cohort |
| Urban/ rural status | Area of residence, urban / rural | Age 12 |
| Household size | Number of individuals in household | Age 12 |
| Wealth index | A composite index of housing quality, access to services and consumer durables subscales, 0-1 | Age 12 |
| Housing quality index | A measure of housing-related comfort, 0-1 | Age 12 |
| Access to services index | A measure of the household’s ability to meet functional requirements of sound shelter, 0-1 | Age 12 |
| Consumer durables index | A measure of the household’s ownership of common household items, 0-1 | Age 12 |
| Debt burden | Whether the household is in serious debt, yes / no | Age 12 |
| Education enrolment | Whether the child is enrolled in school, yes / no | Age 12 |
| Involvement in paid work | Participation in paid work | Age 12 |
| Child’s aspirations for education | The highest level of education the child aspires to reach | Age 12 |
| Caregiver education | The highest level of education achieved by the respondent caregiver | Age 12 |
| Parental aspirations for the child’s education | The highest level of education the parent aspires for the child to reach | Age 12 |
| Caregiver life satisfaction | Cantril ladder life satisfaction rated by the caregiver | Age 12 |

These confounders were chosen based on existing research with Young Lives research on factors shaping trajectories to child marriage and cohabitation in India by Singh and Vennam.^2^

### S5. Data missingness

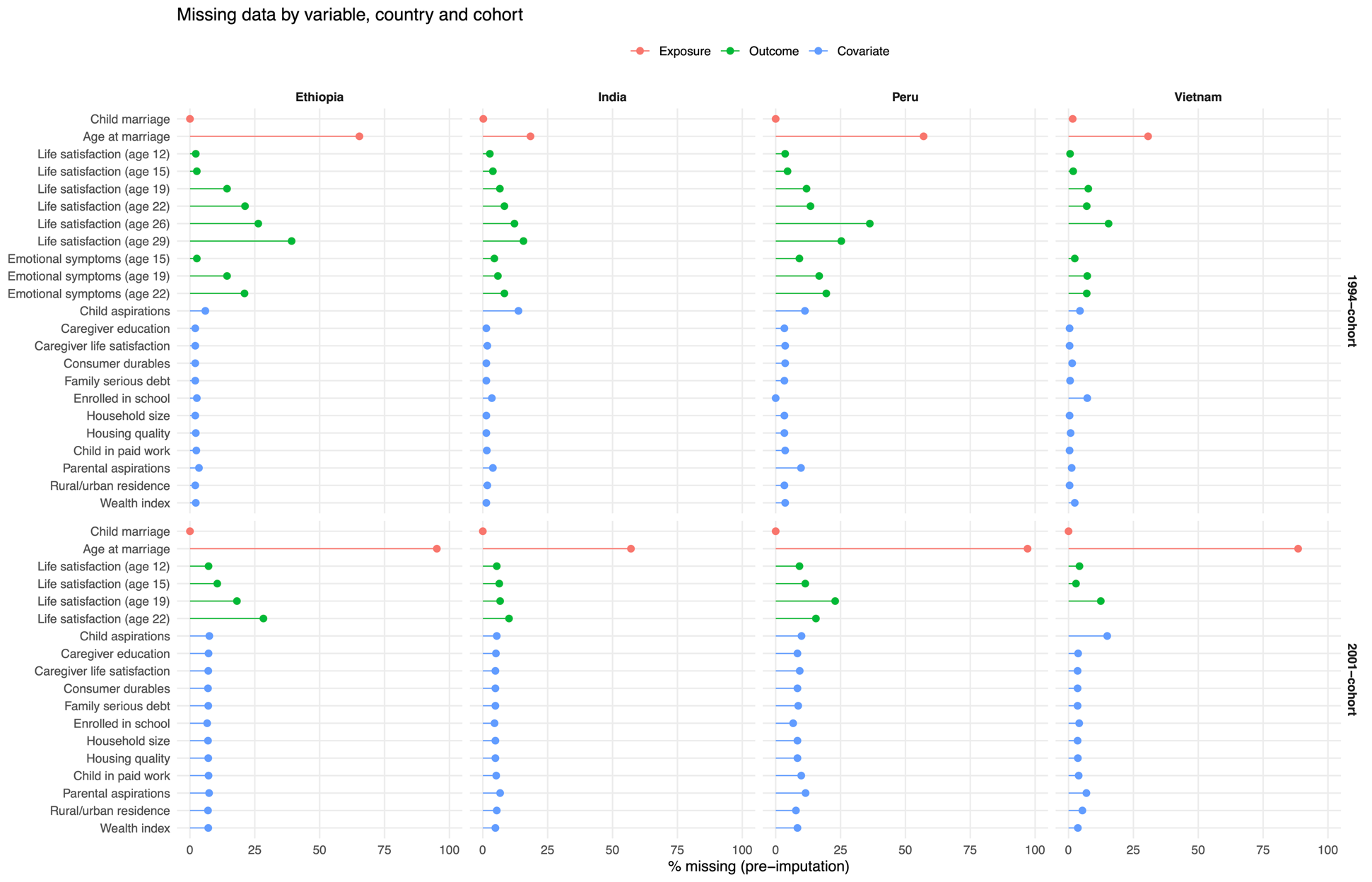

### S6. Rates and mean age of marriage/cohabitation by country and cohort, from the full sample of girls

| **Country** | **Full sample n** | **Child brides, n (%)** | **1994-cohort, n (%)** | **2001-cohort, n (%)** | **Mean age at marriage for child brides, years** |
| --- | --- | --- | --- | --- | --- |
| Ethiopia | 1,440 | 140 (9.7) | 96 (19.6) | 44 (4.6) | 14.3 |
| India | 1,447 | 293 (20.2) | 148 (28.6) | 145 (15.6) | 16.3 |
| Peru | 1,353 | 63 (4.7) | 56 (17.1)* | 7 (0.7) | 16.0 |
| Vietnam | 1,472 | 129 (8.8) | 66 (13.2) | 63 (6.5) | 16.2 |
| Total | 5,712 | 625 (10.9) | 366 (20.0) | 259 (6.7) | 15.8 |

*51 of the 56 girls were in informal unions (cohabited) rather than formal marriage.

### S7. Characteristics of child brides vs non-child brides by country

A) Demographic characteristics

| **Category** | **Variable** | **Ethiopia** | | **India** | | **Peru** | | **Vietnam** | |
| --- | --- | --- | --- | --- | --- | --- | --- | --- | --- |
|  |  | **Non-child-bride (n=1,300)** | **Child bride (n=140)** | **Non-child-bride (n=1,153)** | **Child bride (n=293)** | **Non-child-bride (n=1,300)** | **Child bride (n=140)** | **Non-child-bride (n=1,153)** | **Child bride (n=293)** |
| Sample size (% of cohort total) | 1994-cohort | 394 (80.4%) | 96 (19.6%) | 368 (71.3%) | 148 (28.7%) | 272 (82.9%) | 56  (17.1%) | 433  (86.8%) | 66  (13.2%) |
|  | 2001-cohort | 906 (95.4%) | 44  (4.6%) | 785 (84.4%) | 145 (15.6%) | 1018 (99.3%) | 7  (0.7%) | 910  (93.5%) | 63  (6.5%) |
| Marriage | Age at marriage,  mean (SD) | 19.82 (1.39) | 14.26 (2.92) | 20.86 (2.6) | 16.31 (1.61) | 19.81 (1.84) | 16.05 (1.14) | 21.14 (2.45) | 16.23 (1.1) |
| Outcome | Life satisfaction, mean (SD) | 5.28 (1.91) | 4.64 (1.82) | 4.32 (1.63) | 3.69 (1.6) | 6.49 (1.95) | 6.1 (1.9) | 5.31 (1.7) | 4.55 (1.8) |
| Geography | Rural, n (%) | 750  (57.7%) | 113 (80.7%) | 775 (67.2%) | 256  (87.4%) | 321 (24.9%) | 18  (28.6%) | 1013  (75.4%) | 118  (91.5%) |
|  | Urban, n (%) | 474  (36.5%) | 27  (19.3%) | 324 (28.1%) | 33  (11.3%) | 878 (68.1%) | 45  (71.4%) | 280  (20.8%) | 7  (5.4%) |
| Household characteristics | Birth order, mean (SD) | 2.59 (2.41) | 2.86 (2.64) | 1.11 (1.34) | 1.27 (1.29) | 1.65 (2.01) | 2.24 (2.59) | 0.94 (1.17) | 1.43 (1.8) |
|  | Consumer durables (range 0-1), mean (SD) | 0.27 (0.19) | 0.21 (0.16) | 0.34 (0.19) | 0.24 (0.17) | 0.46 (0.22) | 0.32 (0.2) | 0.58 (0.17) | 0.41 (0.21) |
|  | Household size,  mean (SD) | 6.04 (2.05) | 6.16 (1.89) | 5.02 (1.79) | 4.98 (1.64) | 5.31 (1.85) | 5.95 (2.61) | 4.7 (1.36) | 5.35 (1.68) |
|  | Housing quality  (range 0-1), mean (SD) | 0.34 (0.2) | 0.27 (0.19) | 0.65 (0.26) | 0.57 (0.25) | 0.45 (0.25) | 0.33 (0.21) | 0.75 (0.19) | 0.64 (0.22) |
|  | Wealth index  (range 0-1), mean (SD) | 0.37 (0.19) | 0.29 (0.17) | 0.56 (0.19) | 0.46 (0.15) | 0.57 (0.21) | 0.43 (0.21) | 0.64 (0.16) | 0.48 (0.18) |
|  | Child in paid work, n (%) | 22  (1.7%) | 5  (3.6%) | 14  (1.2%) | 23  (7.8%) | 67  (5.2%) | 15  (23.8%) | 27  (2%) | 15  (11.6%) |
|  | Family has serious debt, n (%) | 437 (33.6%) | 63 (45%) | 526 (45.6%) | 151 (51.5%) | 383 (29.7%) | 17 (27%) | 598 (44.5%) | 71 (55%) |
| Education & aspirations | Enrolled in school, n (%) | 1170  (90%) | 130 (92.9%) | 1057 (91.6%) | 231  (78.8%) | 1199 (92.9%) | 59  (93.7%) | 1241  (92.4%) | 108  (83.7%) |
|  | Child aspirations: post-secondary, n (%) | 867 (66.7%) | 90 (64.3%) | 827 (71.7%) | 141 (48.1%) | 1029 (79.8%) | 46 (73%) | 971 (72.3%) | 55 (42.6%) |
|  | Parental aspirations: post-secondary, n (%) | 972 (74.8%) | 108 (77.1%) | 742 (64.3%) | 102 (34.8%) | 1012 (78.4%) | 42 (66.7%) | 1068 (79.5%) | 61 (47.3%) |
| Caregiver education | None | 488 (37.5%) | 68 (48.6%) | 497 (43.1%) | 205  (70%) | 127 (9.8%) | 10  (15.9%) | 95  (7.1%) | 49  (38%) |
|  | Other | 158 (12.2%) | 29 (20.7%) | 33 (2.9%) | 11 (3.8%) | / | / | / | / |
|  | Primary-secondary | 538 (41.4%) | 42 (30%) | 535 (46.4%) | 75 (25.6%) | 879 (68.1%) | 50 (79.4%) | 1142 (85%) | 77 (59.7%) |
|  | Vocational/ university | 38 (2.9%) | 1 (0.7%) | 36 (3.1%) | 1 (0.3%) | 187 (14.5%) | 3 (4.8%) | 70 (5.2%) | 1 (0.8%) |
| Caregiver life satisfaction | Caregiver life satisfaction  (0-9), mean (SD) | 4.46 (1.55) | 3.99 (1.62) | 3.92 (1.45) | 3.44 (1.38) | 5.48 (1.93) | 4.94 (2.08) | 4.52 (1.47) | 3.97 (1.45) |
| Religion | Ancestor worship | / | / | / | / | / | / | 56 (4.2%) | 2 (1.6%) |
|  | Buddhist | / | / | 8 (0.7%) | 1 (0.3%) | / | / | 93 (6.9%) | 8 (6.2%) |
|  | Cao Dai | / | / | / | / | / | / | 15 (1.1%) | / |
|  | Catholic | 7 (0.5%) | 1 (0.7%) | / | / | 1048 (81.2%) | 50 (79.4%) | / | / |
|  | Christian | / | / | 59 (5.1%) | 13 (4.4%) | / | / | 26 (1.9%) | / |
|  | Evangelist | 1 (0.1%) | / | / | / | 182 (14.1%) | 11 (17.5%) | / | / |
|  | Hindu | / | / | 1006 (87.2%) | 265 (90.4%) | 1 (0.1%) | / | / | / |
|  | Mormon/Other | / | / | / | / | 3 (0.2%) | / | / | / |
|  | Muslim | 214 (16.5%) | 24 (17.1%) | 80 (6.9%) | 13 (4.4%) | / | / | / | / |
|  | None | / | / | / | 1 (0.3%) | 51 (4%) | 1 (1.6%) | 1139 (84.8%) | 118 (91.5%) |
|  | Orthodox | 920 (70.8%) | 100 (71.4%) | / | / | / | / | / | / |
|  | Other | 17 (1.3%) | / | / | / | 4 (0.3%) | 1 (1.6%) | 1 (0.1%) | / |
|  | Protestant | 141 (10.8%) | 15 (10.7%) | / | / | / | / | 12 (0.9%) | 1 (0.8%) |
|  | Sikh | / | / | 1 (0.1%) | / | / | / | / | / |
| Ethnicity | Amhara | 364 (28%) | 36 (25.7%) | / | / | / | / | / | / |
|  | Asian | / | / | / | / | 2 (0.2%) | / | / | / |
|  | Backward Castes | / | / | 518 (44.9%) | 158 (53.9%) | / | / | / | / |
|  | Black | / | / | / | / | 7 (0.5%) | / | / | / |
|  | Dao | / | / | / | / | / | / | 22 (1.6%) | 12 (9.3%) |
|  | Gurage | 114 (8.8%) | 13 (9.3%) | / | / | / | / | / | / |
|  | H'Mong | / | / | / | / | / | / | 51 (3.8%) | 24 (18.6%) |
|  | Kinh | / | / | / | / | / | / | 1210 (90.1%) | 65 (50.4%) |
|  | Mestizo | / | / | / | / | 1185 (91.9%) | 57 (90.5%) | / | / |
|  | Native Amazon | / | / | / | / | 33 (2.6%) | 5 (7.9%) | / | / |
|  | Oromo | 251 (19.3%) | 46 (32.9%) | / | / | / | / | / | / |
|  | Other | / | / | / | / | / | / | 24 (1.8%) | 23 (17.8%) |
|  | Other, Hindu | / | / | 188 (16.3%) | 21 (7.2%) | / | / | / | / |
|  | Other, Muslim | / | / | 65 (5.6%) | 10 (3.4%) | / | / | / | / |
|  | Scheduled Castes | / | / | 220 (19.1%) | 63 (21.5%) | / | / | / | / |
|  | Scheduled Tribes | / | / | 156 (13.5%) | 38 (13%) | / | / | / | / |
|  | Tay | / | / | / | / | / | / | 21 (1.6%) | 1 (0.8%) |
|  | Tigrian | 293 (22.5%) | 21 (15%) | / | / | / | / | / | / |
|  | White | / | / | / | / | 62 (4.8%) | 1 (1.6%) | / | / |
|  | Wolayta | 92 (7.1%) | 5 (3.6%) | / | / | / | / | / | / |

B) Standardised mean differences of demographics

| **Variable** | **Ethiopia** | **India** | **Peru** | **Vietnam** |
| --- | --- | --- | --- | --- |
| Birth order | 0.11 | 0.13 | 0.25 | 0.32 |
| Household size | 0.06 | -0.02 | 0.29 | 0.43 |
| Rural residence | 0.51 | 0.5 | 0.08 | 0.44 |
| Consumer durables index | -0.37 | -0.57 | -0.62 | -0.91 |
| Housing quality index | -0.39 | -0.3 | -0.53 | -0.51 |
| Wealth index | -0.47 | -0.6 | -0.67 | -0.91 |
| Child in paid work | 0.12 | 0.32 | 0.55 | 0.39 |
| Family has serious debt | 0.23 | 0.12 | -0.06 | 0.21 |
| Child enrolled in school | 0.1 | -0.37 | 0.03 | -0.27 |
| Child has aspirations for post-secondary education | -0.05 | -0.49 | -0.16 | -0.63 |
| Caregiver life satisfaction | -0.29 | -0.34 | -0.27 | -0.38 |
| Caregiver has no education | 0.17 | 0.52 | 0.15 | 0.8 |

| SMD | > 0.1 indicates a non-negligible imbalance between the two groups. Multi-category variables such as religion and ethnicity are reported in the previous table of full demographics.

### S8. Post age-12 sensitivity checks excluding girls married/cohabited by age 12

|  | **ETHIOPIA** | | | **INDIA** | | | **PERU^** | | | **VIETNAM^^** | | |
| --- | --- | --- | --- | --- | --- | --- | --- | --- | --- | --- | --- | --- |
|  | **Combined** | **1994-cohort** | **2001-cohort** | **Combined** | **1994-cohort** | **2001-cohort** | **Combined** | **1994-cohort** | **2001-cohort** | **Combined** | **1994-cohort** | **2001-cohort** |
| 1) Life satisfaction at age 19 | | | | | | | | | | | | |
| Adjusted for baseline life satisfaction only | -0.35 (-0.71, 0.01) ** | -0.27 (-0.65, 0.11) | -0.60 (-1.45, 0.25) | -0.57 (-0.79, -0.35) ** | -0.65 (-0.95, -0.36) ** | -0.55 (-0.88, -0.23) ** | -0.12 (-0.60, 0.36) | -0.13 (-0.63, 0.38) | / | -0.76 (-1.08, -0.44) ** | -0.99 (-1.37, -0.61) ** | -0.54 (-1.15, 0.07) ** |
| Fully adjusted | -0.40 (-0.75, -0.05) ** | -0.27 (-0.67, 0.12) | -1.05 (-1.77, -0.34) ** | -0.25 (-0.45, -0.04) ** | -0.38 (-0.70, -0.05) ** | -0.21 (-0.57, 0.14) | -0.02 (-0.54, 0.50) | -0.05 (-0.57, 0.46) | / | -0.32 (-0.70, 0.06) | -0.52 (-0.98, -0.06) ** | -0.23 (-0.77, 0.31) |
| 2) Difference-in-differences estimation from age 12 to 19 | | | | | | | | | | | | |
| Adjusted for baseline life satisfaction only | 0.56 (-0.11, 1.22) | / | / | -0.01 (-0.26, 0.25) | / | / | 0.48 (-0.30, 1.26) | / | / | -0.27 (-0.56, 0.01) ** | / | / |
| Fully adjusted | 0.40 (-0.26, 1.06) | / | / | -0.01 (-0.26, 0.25) | / | / | 0.49 (-0.29, 1.27) | / | / | -0.12 (-0.45, 0.22) | / | / |
| 3) Average life satisfaction across ages 19, 22, 26, and 29 | | | | | | | | | | | | |
| Adjusted for baseline life satisfaction only | -0.28 (-0.51, -0.06) ** | -0.28 (-0.53, -0.03) ** | -0.43 (-0.92, 0.06) ** | -0.44 (-0.58, -0.31) ** | -0.50 (-0.68, -0.31) ** | -0.39 (-0.59, -0.19) ** | -0.08 (-0.38, 0.22) | -0.15 (-0.47, 0.16) | / | -0.66 (-0.90, -0.42) ** | -0.76 (-1.02, -0.49) ** | -0.19 (-1.20, 0.81) |
| Fully adjusted | -0.29 (-0.52, -0.06) ** | -0.31 (-0.56, -0.06) ** | -0.57 (-1.13, -0.01) ** | -0.21 (-0.34, -0.08) ** | -0.26 (-0.44, -0.08) ** | -0.16 (-0.35, 0.03) ** | -0.02 (-0.32, 0.27) | -0.10 (-0.43, 0.24) | / | -0.36 (-0.62, -0.10) ** | -0.52 (-0.83, -0.22) ** | -0.19 (-1.18, 0.79) |
| 4) Trajectories from age 19 to 29 | | | | | | | | | | | | |
| Adjusted for baseline life satisfaction only | / | -0.03 (-0.08, 0.02) | / | / | -0.02 (-0.06, 0.01) | / | / | 0.07 (0.01, 0.12) ** | / | / | 0.05 (-0.02, 0.12) | / |
| Fully adjusted | / | -0.03 (-0.08, 0.02) | / | / | -0.02 (-0.06, 0.01) | / | / | 0.07 (0.01, 0.12) ** | / | / | 0.03 (-0.04, 0.11) | / |
| 5) Emotional symptoms at age 19 | | | | | | | | | | | | |
| Adjusted for baseline life satisfaction only | / | 0.29 (-0.24, 0.82) | / | / | 0.17 (-0.29, 0.63) | / | / | -0.65 (-1.39, 0.09) ** | / | / | 0.49 (-0.00, 0.98) ** | / |
| Fully adjusted | / | 0.12 (-0.38, 0.61) | / | / | -0.01 (-0.48, 0.47) | / | / | -0.80 (-1.67, 0.07) ** | / | / | 0.48 (-0.21, 1.17) | / |
| 6) Average emotional symptoms across ages 19 and 22 | | | | | | | | | | | | |
| Adjusted for baseline life satisfaction only | / | 0.32 (-0.05, 0.68) ** | / | / | 0.11 (-0.23, 0.45) | / | / | -0.49 (-1.18, 0.20) | / | / | 0.38 (0.00, 0.76) ** | / |
| Fully adjusted | / | 0.27 (-0.11, 0.65) | / | / | -0.07 (-0.44, 0.29) | / | / | -0.62 (-1.32, 0.08) ** | / | / | 0.43 (-0.03, 0.89) ** | / |

** p<0·05, * p<0.1

### S9. Propensity score matching (PSM) sample sizes

The two cohorts were matched separately using propensity score matching with 1:1 optimal matching, but in Peru 2001-cohort was not matched due to very few child brides (n=7), hence only the 1994-cohort models were run.

The sample sizes of the matched cohorts are as follows:

- Ethiopia: 96 child brides and 96 non-child-brides in 1994-cohort, 44 child brides and 44 non-child-brides in 2001-cohort
- India: 148 child brides and 148 non-child-brides in 1994-cohort, 145 child brides and 145 non-child-brides in 2001-cohort
- Peru: 56 child brides and 56 non-child-brides in 1994-cohort
- Vietnam: 66 child brides and 66 non-child-brides in 1994-cohort, 63 child brides and 63 non-child-brides in 2001-cohort

### S10. Difference-in-differences (DiD) estimation of life satisfaction change from age 12 to 19 with combined cohorts

|  | **ETHIOPIA**  n=1,263 (CB=137) | **INDIA**  n=1,446 (CB=293) | **PERU**  n=1,308 (CB=63) | **VIETNAM**  n=1,459 (CB=129) |
| --- | --- | --- | --- | --- |
| Adjusted for baseline life satisfaction only | 0.55 * (-0.07, 1.17) | -0.00 (-0.26, 0.26) | -0.26 * (-0.55, 0.03) | -0.26 * (-0.55, -0.03) |
| Fully adjusted | 0.38 (-0.23, 0.99) | -0.00 (-0.26, 0.26) | -0.10 (-0.44, 0.23) | -0.10 (-0.44, 0.23) |
| PSM | -0.51 (-1.12, 0.10) | -0.25 (-0.62, 0.12) | 0.25 (-0.84, 1.34) | -0.42 * (-0.89, 0.04) |

* p<0.1

Note: Parallel trends test was only possible with the 2001-cohort as prior trends (at age 8) was not available for the 1994-cohort.

### S11. Life satisfaction trajectories across ages 19-29 by child marriage status in the 1994-cohort only across countries, adjusted for baseline life satisfaction and covariates

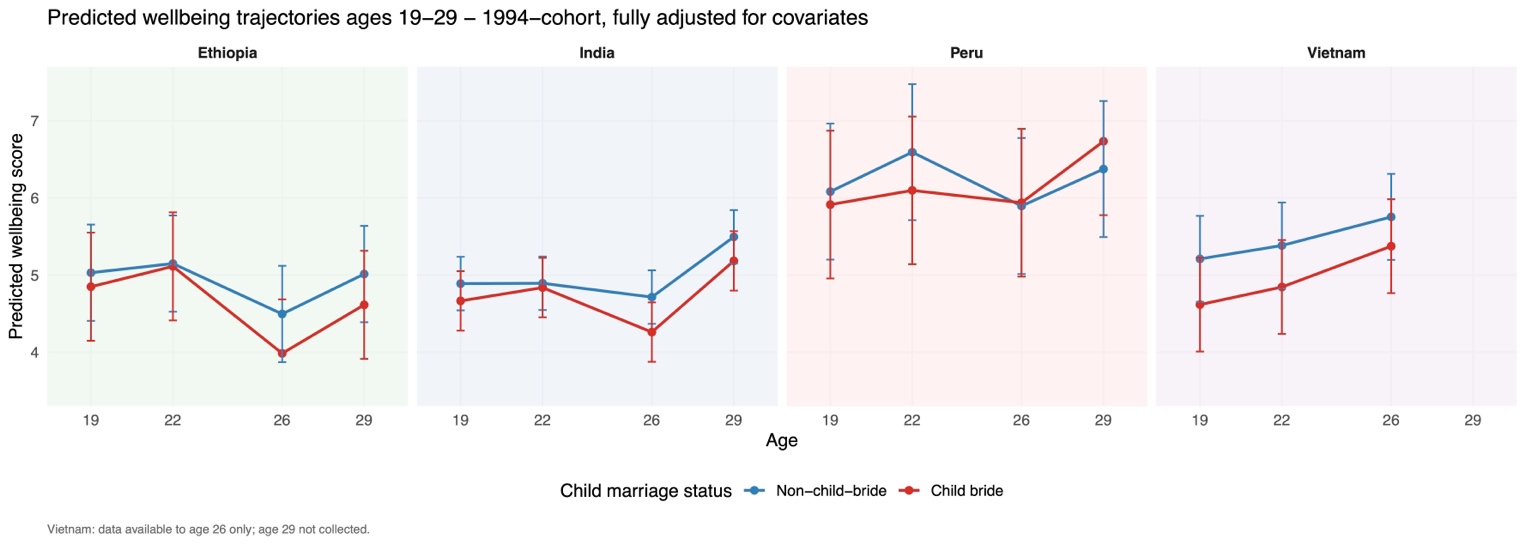

### S12. Age of marriage as exposure

A) Sample sizes

| Country | Total sample | 1994-cohort CB total | 1994-cohort CB married <=15 | 1994-cohort CB married 16-17 | 2001-cohort CB total | 2001-cohort CB married <=15 | 2001-cohort CB married 16-17 |
| --- | --- | --- | --- | --- | --- | --- | --- |
| Ethiopia | 99 | 55 | 28 | 27 | 44 | 23 | 21 |
| India | 293 | 148 | 75 | 73 | 145 | 59 | 86 |
| Peru | 63 | 56 | 12 | 44 | 7 | 3 | 4 |
| Vietnam | 115 | 66 | 12 | 54 | 49 | 11 | 38 |

B) Summary of results

Among Ethiopia’s 1994-cohort child brides, those who married/cohabited later had higher predicted emotional symptoms at age 19 (0.23[0.04, 0.43]) and across ages 19-22 (0.19[-0.01, 0.39]), however, these associations attenuated after excluding individuals married by age 12 (n=26). We found no association between age of marriage/cohabitation and life satisfaction in this sensitivity check nor in propensity score matched samples. Differences-in-differences estimation with combined cohorts found no differences in change in life satisfaction nor emotional symptoms between girls married before 15 years compared to those married 16-17 years in all specifications.

C) Unmatched and matched models

Age of marriage was a continuous predictor in all models except the DiD estimation, where those married by 15 years were compared to those married at 16-17. Matched analyses were conducted with combined cohorts in Ethiopia and India predicting life satisfaction at 19 and across 19-29 only as sample sizes were too small in Peru and Vietnam and for other models. In Peru’s 2001-cohort there are too few child brides (n=7) to run any model, hence there were no results for 2001-cohort or combined cohort. In Vietnam age 29 is not available for 1994-cohort, hence Model 2 is up to age 26 only. For 2001-cohort only age 19 data available, thus equivalent to Model 1.

|  | **ETHIOPIA** | | | **INDIA** | | | **PERU** | | | **VIETNAM** | | |
| --- | --- | --- | --- | --- | --- | --- | --- | --- | --- | --- | --- | --- |
|  | **Combined** | **1994-cohort** | **2001-cohort** | **Combined** | **1994-cohort** | **2001-cohort** | **Combined** | **1994-cohort** | **2001-cohort** | **Combined** | **1994-cohort** | **2001-cohort** |
| 1) Life satisfaction at age 19 | | | | | | | | | | | | |
| Adjusted for baseline life satisfaction only | -0.13 (-0.21, -0.05) ** | -0.17 (-0.30, -0.04) ** | -0.10 (-0.21, 0.00) ** | -0.02 (-0.15, 0.11) | -0.02 (-0.17, 0.14) | -0.01 (-0.23, 0.20) | / | -0.37 (-0.97, 0.23) | / | 0.04 (-0.20, 0.29) | 0.11 (-0.30, 0.51) | -0.09 (-0.58, 0.40) |
| Fully adjusted | -0.04 (-0.16, 0.08) | -0.10 (-0.32, 0.13) | -0.08 (-0.28, 0.12) | -0.04 (-0.19, 0.12) | -0.07 (-0.22, 0.09) | 0.03 (-0.22, 0.29) | / | -0.27 (-1.04, 0.49) | / | 0.01 (-0.29, 0.30) | -0.10 (-0.82, 0.62) | -0.35 (-1.32, 0.62) |
| PSM | 0.22 (-1.16, 1.59) | / | / | 0.02 (-0.45, 0.49) | / | / | / | / | / | / | / | / |
| 2) Difference-in-differences estimation comparing <=15 vs 16-17 groups in life satisfaction change from age 12 to 19 (combined cohorts only) | | | | | | | | | | | | |
| Adjusted for baseline life satisfaction only | 0.17 (-1.23, 1.57) | / | / | -0.03 (-0.67, 0.62) | / | / | 0.30 (-0.95, 1.55) | / | / | 0.08 (-0.95, 1.11) | / | / |
| Fully adjusted | 0.09 (-1.34, 1.53) | / | / | -0.03 (-0.67, 0.62) | / | / | 0.30 (-0.95, 1.55) | / | / | 0.50 (-1.15, 2.15) | / | / |
| 3) Average life satisfaction across ages 19, 22, 26, and 29 | | | | | | | | | | | | |
| Adjusted for baseline life satisfaction only | -0.06 (-0.13, 0.02) | -0.07 (-0.18, 0.04) | -0.05 (-0.16, 0.06) | 0.04 (-0.03, 0.12) | 0.08 (-0.02, 0.17) | -0.03 (-0.14, 0.09) | / | -0.24 (-0.56, 0.08) | / | -0.04 (-0.26, 0.18) | -0.02 (-0.24, 0.20) | / |
| Fully adjusted | 0.03 (-0.07, 0.13) | -0.05 (-0.18, 0.09) | 0.04 (-0.23, 0.30) | 0.02 (-0.05, 0.09) | 0.02 (-0.07, 0.12) | -0.01 (-0.12, 0.11) | / | -0.18 (-0.63, 0.27) | / | 0.04 (-0.24, 0.32) | 0.02 (-0.33, 0.37) | / |
| PSM | 0.38 (-0.32, 1.08) | / | / | 0.08 (-0.20, 0.36) | / | / | / | / | / | / | / | / |
| 4) Trajectories from age 19 to 29 (1994-cohort only) | | | | | | | | | | | | |
| Adjusted for baseline life satisfaction only | / | 0.13 (-0.64, 0.89) | / | / | -0.14 (-0.54, 0.26) | / | / | 0.92 (-0.16, 1.99) ** | / | / | -0.68 (-1.54, 0.19) | / |
| Fully adjusted | / | 0.48 (-0.42, 1.38) | / | / | 0.05 (-0.35, 0.46) | / | / | 0.80 (-0.46, 2.07) | / | / | -0.66 (-1.95, 0.63) | / |
| 5) Emotional symptoms at age 19 (1994-cohort only) | | | | | | | | | | | | |
| Adjusted for baseline life satisfaction only | / | 0.02 (-0.11, 0.16) | / | / | -0.07 (-0.26, 0.12) | / | / | -0.23 (-0.91, 0.45) | / | / | 0.08 (-0.23, 0.39) | / |
| Fully adjusted | / | 0.23 (0.04, 0.43) ** | / | / | -0.01 (-0.23, 0.21) | / | / | -0.13 (-1.25, 0.99) | / | / | -0.03 (-0.88, 0.83) | / |
| 6) Average emotional symptoms across ages 19 and 22 (1994-cohort only) | | | | | | | | | | | | |
| Adjusted for baseline life satisfaction only | / | 0.03 (-0.10, 0.17) | / | / | -0.03 (-0.22, 0.15) | / | / | -0.27 (-0.79, 0.26) | / | / | 0.04 (-0.25, 0.34) | / |
| Fully adjusted | / | 0.19 (-0.01, 0.39) ** | / | / | 0.00 (-0.19, 0.20) | / | / | -0.23 (-1.07, 0.60) | / | / | -0.04 (-0.58, 0.50) | / |

** p<0·05, * p<0.1

D) Post age-12 sensitivity check: excluding girls married/cohabited by age 12

As a sensitivity check, after removing girls married/cohabited by age 12, there remained 73 child brides in Ethiopia, 280 in India, 55 in Peru (1994-cohort only), and 65 in Vietnam.

Models were the same as above, except in Ethiopia 2001-cohort there were too few girls to run fully adjusted models.

|  | **ETHIOPIA** | | | **INDIA** | | | **PERU^** | | | **VIETNAM^^** | | |
| --- | --- | --- | --- | --- | --- | --- | --- | --- | --- | --- | --- | --- |
|  | **Combined** | **1994-cohort** | **2001-cohort** | **Combined** | **1994-cohort** | **2001-cohort** | **Combined** | **1994-cohort** | **2001-cohort** | **Combined** | **1994-cohort** | **2001-cohort** |
| 1) Life satisfaction at age 19 | | | | | | | | | | | | |
| Adjusted for baseline life satisfaction only | -0.07 (-0.29, 0.14) | -0.01 (-0.34, 0.31) | -0.21 (-0.50, 0.07) | -0.03 (-0.19, 0.12) | 0.02 (-0.14, 0.19) | -0.11 (-0.42, 0.20) | / | -0.52 (-1.18, 0.15) | / | 0.13 (-0.12, 0.38) | 0.29 (0.05, 0.53) ** | -0.09 (-0.58, 0.40) |
| Fully adjusted | -0.03 (-0.51, 0.44) | -0.18 (-0.72, 0.35) | N/A | -0.06 (-0.22, 0.10) | 0.00 (-0.17, 0.18) | -0.15 (-0.48, 0.19) | / | -0.43 (-1.43, 0.58) | / | 0.08 (-0.28, 0.44) | 0.20 (-0.30, 0.69) | -0.35 (-1.32, 0.62) |
| 2) Average life satisfaction across ages 19, 22, 26, and 29 | | | | | | | | | | | | |
| Adjusted for baseline life satisfaction only | -0.03 (-0.24, 0.17) | -0.02 (-0.25, 0.21) | -0.13 (-0.60, 0.33) | 0.01 (-0.08, 0.10) | 0.05 (-0.06, 0.16) | -0.10 (-0.25, 0.05) |  | -0.23 (-0.62, 0.16) | / | 0.02 (-0.24, 0.27) | 0.07 (-0.20, 0.34) | / |
| Fully adjusted | 0.03 (-0.23, 0.29) | -0.01 (-0.30, 0.29) | N/A | -0.01 (-0.09, 0.08) | 0.02 (-0.09, 0.12) | -0.10 (-0.25, 0.06) |  | -0.16 (-0.66, 0.33) | / | 0.15 (-0.21, 0.50) | 0.24 (-0.20, 0.68) | / |
| 3) Trajectories from age 19 to 29 | | | | | | | | | | | | |
| Adjusted for baseline life satisfaction only | / | -0.01 (-0.13, 0.12) | / | / | 0.00 (-0.05, 0.06) | / | / | -0.10 (-0.23, 0.03) | / | / | 0.18 (-0.02, 0.38) ** | / |
| Fully adjusted | / | 0.00 (-0.12, 0.12) | / | / | 0.00 (-0.05, 0.06) | / | / | -0.10 (-0.23, 0.03) | / | / | 0.10 (-0.15, 0.36) | / |
| 4) Emotional symptoms at age 19 | | | | | | | | | | | | |
| Adjusted for baseline life satisfaction only | / | -0.28 (-0.88, 0.32) | / | / | 0.03 (-0.22, 0.29) | / | / | -0.37 (-1.37, 0.63) | / | / | 0.09 (-0.35, 0.52) | / |
| Fully adjusted | / | 0.02 (-0.84, 0.88) | / | / | 0.07 (-0.19, 0.33) | / | / | -0.01 (-1.31, 1.28) | / | / | 0.04 (-1.17, 1.25) | / |
| 5) Average emotional symptoms across ages 19 and 22 | | | | | | | | | | | | |
| Adjusted for baseline life satisfaction only | / | -0.04 (-0.33, 0.25) | / | / | 0.01 (-0.21, 0.24) | / | / | -0.51 (-1.13, 0.11) | / | / | 0.11 (-0.25, 0.46) | / |
| Fully adjusted | / | 0.15 (-0.27, 0.57) | / | / | 0.06 (-0.17, 0.29) | / | / | -0.35 (-1.28, 0.59) | / | / | -0.01 (-0.70, 0.69) | / |

** p<0·05, * p<0.1

### S13. Strobe checklist

STROBE Statement—checklist of items that should be included in reports of observational studies

|  | Item No. | Recommendation | Page  No. | Relevant text from manuscript | |
| --- | --- | --- | --- | --- | --- |
| Title and abstract | 1 | (*a*) Indicate the study’s design with a commonly used term in the title or the abstract | 1 | Title | |
|  |  | (*b*) Provide in the abstract an informative and balanced summary of what was done and what was found | 2 | Abstract | |
| Introduction | | | |  | |
| Background/rationale | 2 | Explain the scientific background and rationale for the investigation being reported | 5-6 | Introduction | |
| Objectives | 3 | State specific objectives, including any prespecified hypotheses | 6 | Introduction | |
| Methods | | | |  | |
| Study design | 4 | Present key elements of study design early in the paper | 6 | Introduction, Methods - Participants | |
| Setting | 5 | Describe the setting, locations, and relevant dates, including periods of recruitment, exposure, follow-up, and data collection | 6 | Methods - Participants | |
| Participants | 6 | (*a*) *Cohort study*—Give the eligibility criteria, and the sources and methods of selection of participants. Describe methods of follow-up | 6 | Methods - Participants, Supplement S1 | |
|  |  | (*b*) *Cohort study*—For matched studies, give matching criteria and number of exposed and unexposed | 7 | Methods – Statistical analysis, Supplement S9 | |
| Variables | 7 | Clearly define all outcomes, exposures, predictors, potential confounders, and effect modifiers. Give diagnostic criteria, if applicable | 7 | Methods – Measures | |
| Data sources/ measurement | 8* | For each variable of interest, give sources of data and details of methods of assessment (measurement). Describe comparability of assessment methods if there is more than one group | 7-9 | Methods – Measures, Supplement S2 | |
| Bias | 9 | Describe any efforts to address potential sources of bias | 8-9 | Methods – Statistical analysis, Supplement S8 | |
| Study size | 10 | Explain how the study size was arrived at | Supplement S1 | Supplement S1 flowchart | |
| Quantitative variables | 11 | Explain how quantitative variables were handled in the analyses. If applicable, describe which groupings were chosen and why | 6-7 | | Methods – Statistical analysis |
| Statistical methods | 12 | (*a*) Describe all statistical methods, including those used to control for confounding | 7 | | Methods – Statistical analysis |
|  |  | (*b*) Describe any methods used to examine subgroups and interactions | 7 | | Methods – Statistical analysis |
|  |  | (*c*) Explain how missing data were addressed | 7-8 | | Methods – Statistical analysis |
|  |  | (*d*) *Cohort study*—If applicable, explain how loss to follow-up was addressed |  | | Methods – Statistical analysis |
|  |  | (*e*) Describe any sensitivity analyses | 7-9 | | Methods – Statistical analysis |
| Results | | | | | |
| Participants | 13* | (a) Report numbers of individuals at each stage of study—eg numbers potentially eligible, examined for eligibility, confirmed eligible, included in the study, completing follow-up, and analysed | Results, Supplement S1 | | Supplement S1 |
|  |  | (b) Give reasons for non-participation at each stage | Supplement S1 | | Supplement S1 |
|  |  | (c) Consider use of a flow diagram | Supplement S1 | | Supplement S1 flowchart |
| Descriptive data | 14* | (a) Give characteristics of study participants (eg demographic, clinical, social) and information on exposures and potential confounders | Table 1, Supplement S2 | | Table 1, Supplement S2 |
|  |  | (b) Indicate number of participants with missing data for each variable of interest | Supplement S5 | | Supplement S5 |
|  |  | (c) *Cohort study*—Summarise follow-up time (eg, average and total amount) | 9, Figure 1, Supplement S1 | | Methods – Participants, Supplement S1 |
| Outcome data | 15* | *Cohort study*—Report numbers of outcome events or summary measures over time | 6-7 | | Methods – Measures |
| Main results | 16 | (*a*) Give unadjusted estimates and, if applicable, confounder-adjusted estimates and their precision (eg, 95% confidence interval). Make clear which confounders were adjusted for and why they were included | 10-11 | | Results |
|  |  | (*b*) Report category boundaries when continuous variables were categorized | 10-11 | | Results |
|  |  | (*c*) If relevant, consider translating estimates of relative risk into absolute risk for a meaningful time period |  | |  |
| Other analyses | 17 | Report other analyses done—eg analyses of subgroups and interactions, and sensitivity analyses | 10-11, Table 2, Supplement S8 | | Table 2 by cohort, Supplement S8 post age-12 sensitivity |
| Discussion | | | | | |
| Key results | 18 | Summarise key results with reference to study objectives | 12 | | Discussion |
| Limitations | 19 | Discuss limitations of the study, taking into account sources of potential bias or imprecision. Discuss both direction and magnitude of any potential bias | 13 | | Discussion |
| Interpretation | 20 | Give a cautious overall interpretation of results considering objectives, limitations, multiplicity of analyses, results from similar studies, and other relevant evidence | 12-14 | | Discussion |
| Generalisability | 21 | Discuss the generalisability (external validity) of the study results | 12-14 | | Discussion |
| Other information | |  | | | |
| Funding | 22 | Give the source of funding and the role of the funders for the present study and, if applicable, for the original study on which the present article is based | 15 | | Acknowledgements |

### Bibliography

1	Favara M, De Los Ángeles Molina M, Sánchez A, et al. Cohort Profile Update: The Young Lives study. Int J Epidemiol 2026; 55: dyag087.

2	Singh R, Vennam U. Factors Shaping Trajectories to Child and Early Marriage: Evidence from Young Lives in India. 2016 https://www.younglives.org.uk/sites/default/files/migrated/YL-WP149-Trajectories%20to%20early%20Marriage.pdf (accessed Feb 17, 2025).
